# Associations of genetic scores for birth weight with newborn size and later Anthropometric traits and cardiometabolic risk markers in South Asians

**DOI:** 10.1101/2021.04.16.21254284

**Authors:** Suraj S Nongmaithem, Robin N Beaumont, Akshay Dedaniya, Andrew R Wood, Babatunji-William Ogunkolade, Zahid Hassan, Ghattu V Krishnaveni, Kalyanaraman Kumaran, Ramesh D Potdar, Sirajul A Sahariah, Murali Krishna, Chiara Di Gravio, Inder D Mali, Alagu Sankareswaran, Akhtar Hussain, Biswajit W Bhowmik, Abdul Kalam A Khan, Bridget A Knight, Timothy M Frayling, Sarah Finer, Caroline HD Fall, Chittaranjan S Yajnik, Rachel M Freathy, Graham A Hitman, Giriraj R Chandak

## Abstract

We recently reported genetic variants associated with birth weight and their effect on future cardiometabolic risk in Europeans. Despite a higher burden of low birth weight and cardiometabolic disorders, such studies are lacking in South Asians. We generated fetal and maternal genetic scores (fGS and mGS) from 196 birth weight-associated variants identified in Europeans and conducted association analysis with various birth measures and serially measured anthropometric and cardiometabolic traits from seven Indian and Bangladeshi cohorts. Although fGS and mGS were comparable to Europeans, birth weight was substantially smaller suggesting strong environmental constraints on fetal growth in South Asians. Birth weight increased by 50.7g and 33.6g per standard deviation fGS (P=9.1×10^−11^) and mGS (P=0.003) in South Asians. The fGS was further associated with childhood body size and head circumference, fasting glucose, and triglycerides in adults (P<0.01). Our study supports a common genetic mechanism partly explaining associations between early development and later cardiometabolic health in different populations, despite phenotypic and environmental differences.

## INTRODUCTION

Size at birth is a summary measure for intrauterine nutrition, growth and development^1, 2^. It is influenced by genetic and environmental factors, and in clinical practice helps predict neonatal wellbeing^3, 4^. Several longitudinal population-based studies both in higher and lower-middle-income countries including India have demonstrated a correlation between birth size (both small and large) and future risk of cardiometabolic diseases including type 2 diabetes (T2D) and cardiovascular disease (CVD)^1, 2, 5, 6, 7, 8^. This led to the ‘Fetal Programming’ or Developmental Origins of Health and Disease (DOHaD) hypothesis which proposes that the fetus’s environment (meaning maternal diet, workload, smoking, etc) drives its growth and also affects the development of metabolic organs, setting up later risk of disease^1, 2^. South Asians living in the Indian sub-continent have a high prevalence of low birth weight as well as a rising epidemic of T2D and CVD. Moreover, they develop these conditions at a younger age and a lower body mass index (BMI) than Europeans^9^. Fetal growth is influenced by various fetal and maternal factors including genetic effects, maternal size, and maternal health and nutrition^4^. Understanding the genetic determinants of the neonatal size and their association with later phenotypes may provide important insights into mechanisms of how fetal growth and development relate to later risk of cardiometabolic diseases especially in different ethnic groups with different environmental exposures.

Large-scale genome-wide association studies (GWASs), mostly in individuals of European ancestry, including participants from the Early Growth Genetics (EGG) consortium and the UK Biobank (UKBB) have identified several genetic variants associated with birth weight^10, 11, 12, 13, 14^. These genetic associations include (i) direct effects, where the fetus’s own genotype influences its birth weight, (ii) indirect effects of the maternal genetic factors which influence birth weight via the intrauterine environment, and (iii) those which have a combination of direct fetal and indirect maternal effects^10, 14^. The latest study by Warrington and colleagues updated the number of genetic variants associated with birth weight to 209 single nucleotide polymorphisms (SNPs) at 190 independent loci^14^. It further partitioned the genetic effects on birth weight into fetal and maternal effects and demonstrated their association with various cardiometabolic traits. Based on the observations that fetal genetic score for birth weight is negatively associated with adult blood pressure (BP), lipids, glucose and insulin levels, and insulin resistance, Warrington et al concluded that common genetic variants contribute to the observed associations between lower birth weight and later cardiometabolic disease. This is something akin to the ‘Fetal Insulin Hypothesis’ first set out by Hatterseley et al^15^. Despite a higher burden of low birth weight and cardio-metabolic disorders, such large-scale studies are lacking in South Asians.

The dual burden of low birth weight and cardiometabolic diseases, and the fact that South Asians, especially those living in lower and middle income countries are not well represented in the majority of GWAS studies to date demands investigating genetic variants associated with their fetal development, and how they relate to the later cardiometabolic traits^16, 17, 18^. Here, we investigate associations of the genetic score derived from the study by Warrington et al with birth size in ∼1900 mother offspring pairs from a number of South Asian birth cohorts in India, Bangladesh and UK. Available data from these cohorts provided us a unique opportunity to investigate associations of the birth weight genetic score with body size/composition and cardiometabolic risk factors among children, adolescents and adults. Overall, the study has tried to answer two questions: (1) are fetal and maternal genetic scores related to newborn size in South Asians in the same way as in Europeans? and (2) do the genetic scores related to birth weight influence cardiometabolic risk in a direction that would support a genetic contribution to the birth weight-cardiometabolic disease link in the South Asian population?

## RESULTS

### Clinical and demographic characteristics of study participants

Newborn measurements, maternal details and phenotypes at different follow-up stages are shown in table 1 and supplementary tables S1A, S1B, S2A and S2B. The mean birth weight of term babies in different cohorts ranged between 2.64 and 3.12 kg. Babies born in India [Pune Maternal Nutrition Study (PMNS; n=515), Parthenon Study (PS; n = 511), Mumbai Maternal Nutrition Project (MMNP; n = 466), Mysore Birth Records Cohort (MBRC; n = 684)] and in Bangladesh [GIFTS work package 2 (WP2; n = 53) and GIFTS work package 3 (WP3; n = 314) cohorts] were comparatively smaller, whereas Bangladeshi babies from the London UK Bangladeshi cohort (UK-Bang; n = 150) and the UK Biobank South Asian component (n=2732) were relatively larger. The birth weight was much higher in the European babies as we observed in the EFSOCH (n=674) (Table 1). Boys were bigger than girls across all the cohorts. In contrast, sum of skin-fold thickness, a measure of adiposity, was greater in girls. Amongst all the cohorts, PMNS mothers living in rural India were the thinnest (mean BMI = 18.0 kg/m^2^) whereas Bangladeshi mothers living in the UK (UK-Bang) were the heaviest (mean BMI = 26.2 kg/m^2^). Mean BMI in the mothers from the other cohorts were in the normal range, between 20.3 to 23.6 kg/m^2^. The percentage of mothers with gestational diabetes mellitus (GDM) was higher in the Bangladeshi cohorts (UK-Bang = 50%, WP2 = 24.5% and WP3 = 25.8%), whereas, in the Indian cohorts, it was 0.6%, 6.1% and 6.9% in PMNS, PS and MMNP respectively. The UK-Bang cohort was positively selected to have higher rates of GDM than the underlying population, but the high rates of GDM in the Bangladeshi Dhaka WP2 and WP3 cohorts represent the high rates of GDM in the community. The mothers of MBRC individuals (born 1934-1966) were not tested for diabetes.

**Table 1.** Maternal and newborn details in the study cohorts, and fetal and maternal genetic scores for the study cohorts and European cohorts.

| Traits | PMNS<br>(N=515) | PS<br>(N=511) | MMNP<br>(N=466) | MBRC<br>(N=684) | Dhaka-WP2<br>(N=53) | Dhaka-WP3<br>(N=314) | UK-Bang<br>(N=150) | UKBB-SAS*<br>(N=2732) | EFSOCH*<br>(N=674) |
| --- | --- | --- | --- | --- | --- | --- | --- | --- | --- |
| Birthweight (kg) | 2.68 (0.34) | 2.91 (0.41) | 2.64 (0.37) | 2.76 (0.42) | 2.90 (0.38) | 2.84 (0.42) | 3.12 (0.45) | 3.10 (0.68) | 3.52 (0.47) |
| Birth length (cm) | 47.8 (1.97) | 48.8 (2.11) | 48.2 (2.26) | 48.0 (2.95) | 46.2 (2.56) | 49.6 (2.60) | 46.6 (2.03) | NA | 50.3 (2.12) |
| Ponderal index (kg/m<sup>3</sup>) | 24.5 (2.44) | 25.0 (2.75) | 23.6 (2.60) | 25.3 (4.85) | 29.5 (4.42) | 23.3 (3.50) | 28.9 (4.27) | NA | 27.7 (2.58) |
| Head circumference (cm) | 33.1 (1.24) | 33.9 (1.28) | 33.2 (1.20) | 35.6 (1.58) | 33.4 (1.39) | 33.0 (2.40) | 33.6 (1.31) | NA | 35.2 (1.26) |
| Chest circumference (cm) | 31.2 (1.59) | 32.0 (1.64) | 30.9 (1.75) | NA | NA | NA | 33.4 (1.97) | NA | 34.2 (1.86) |
| Abdomen circumference (cm) | 28.7 (1.91) | 30.0 (1.92) | 28.4 (2.08) | NA | NA | NA | 31.4 (2.56) | NA | NA |
| Mid-upper arm circumference (cm) | 9.7 (0.88) | 10.4 (0.92) | 9.7 (0.82) | NA | 9.9 (0.71) | 10.2 (2.09) | 10.9 (2.13) | NA | 11.1 (0.90) |
| Triceps skinfold (mm) | 4.3 (0.87) | 4.3 (0.90) | 4.2 (1.05) | NA | NA | NA | 5.0 (1.93) | NA | 4.86 (1.08) |
| Subscapular skinfold (mm) | 4.2 (0.89) | 4.5 (0.91) | 4.2 (0.99) | NA | NA | NA | 5.3 (1.87) | NA | 4.87 (1.08) |
| Gestational age (weeks) | 39.0 (1.06) | 39.5 (1.14) | 39.3 (1.17) | NA | 40.3 (1.17) | 39.2 (1.53) | 40.0 (3.44) | NA | 40.1 (1.22) |
| Maternal Age (years) | 21.4 (3.56) | 23.8 (4.24) | 24.8 (3.83) | NA | 19.9 (2.45) | 22.7 (4.29) | 29.7 (5.40) | NA | 30.5 (5.19) |
| Maternal Height (cm) | 152.1(4.9) | 154.5(5.4) | 151.3(5.4) | NA | 151.1 (5.8) | 150.9 (5.7) | 156.0 (5.8) | NA | 165.0 (6.3) |
| Maternal BMI (kg/m<sup>2</sup>) | 18.0 (1.9) | 23.6 (3.55) | 20.3 (3.67) | NA | 20.6 (3.40) | 22.7 (4.03) | 26.2 (4.34) | NA | 24.0 (4.34) |
| Maternal GDM status [n (%)] | 3 (0.6) | 31 (6.1) | 32 (6.9) | NA | 13 (24.5) | 81 (25.8) | 75 (50.0) | NA | NA |
| Year of birth | 1994-95 | 1998-99 | 2006-12 | 1934-66 | 2011-12 | 2015-16 | 2011-15 | 1934-70 | 2000-04 |
| Fetal Genetic Score | 191.0 (9.0) | 191.0 (9.6) | 189.0 (9.4) | 189.0 (9.6) | 191.0 ( 8.1) | 188.0 (9.4) | 188.0 (9.3) | 192.0 (9.9) | 192.0 (9.8) |
| Maternal Genetic Score | 215.0 (10.3) | 215.0 (10.4) | 215.0 (10.5) | NA | 218.0 (10.2) | 217.0 (10.2) | 216.0 (9.3) | NA | 214.0 (10.8) |

All values are mean (SD); N, subjects included in this study; SD, standard deviation; GDM, Gestational diabetes mellitus; PMNS, Pune Maternal Nutrition Study; PS, Parthenon Study; MMNP, Mumbai Maternal Nutrition Project; MBRC, Mysore Birth Records Cohort; Dhaka-WP2, Work Package 2 of GIFTS; Dhaka-WP3, Work Package 3 of GIFTS; UK-Bang, London UK Bangladeshi cohort; UKBB-SAS, UK Biobank South Asian component; EFSOCH, The Exeter Family Study of Childhood Health study; \* Not used for meta-analysis. Fetal and maternal genetic scores were calculated from 196 birthweight-associated variants in children and mothers , respectively.

### Selection of genetic variants, comparison of the effect allele frequency, and calculation of weighted genetic scores

The scheme for selecting SNPs for the calculation of birth weight genetic score is shown in figure 1. Of the 205 autosomal SNPs reported as associated with birth weight by Warrington and colleagues, nine were excluded due to either being missing or having an imputation info score less than 0.4 in at least one of the cohorts. The weighted fetal genetic score (fGS) and weighted maternal genetic score (mGS) were calculated in each cohort from the remaining 196 SNPs using the structural equation modelling (SEM) adjusted fetal and maternal effects respectively from Warrington NM et al. as weights (See Methods)^14^. The SEM estimates associations of both maternal and fetal scores with birth weight while accounting for the relationship between fetal and maternal genotypes, thereby producing independent estimates of the fetal and maternal genetic effects on birth weight. Mean fGS and mGS scores for all South Asian cohorts were similar (range = 188 – 192 for fGS and 214-218 for mGS) (Table 1). Effect allele frequencies (EAFs) of the 196 SNPs were similar in all 7 South Asian cohorts, with the exception of only two outliers, one each in the MBRC (rs2306547) and GIFTS (rs9851257) cohorts (Supplementary Figure S1A). Although, the EAFs at several SNPs varied considerably between South Asians and the EGG/UK Biobank subjects (Supplementary Figure S1B), both fGS and mGS scores in South Asians cohorts combined were similar to that in an European cohort, The Exeter Family Study of Childhood Health (EFSOCH) [(fGS (mean, SD) - 189.8 (9.4) in South Asians vs 192.0 (9.8) in EFSOCH]; [mGS (mean, SD) - 215.5 (10.3) in South Asians vs. 214.0 (10.8) in EFSOCH] (Table 1).

**Figure 1.**
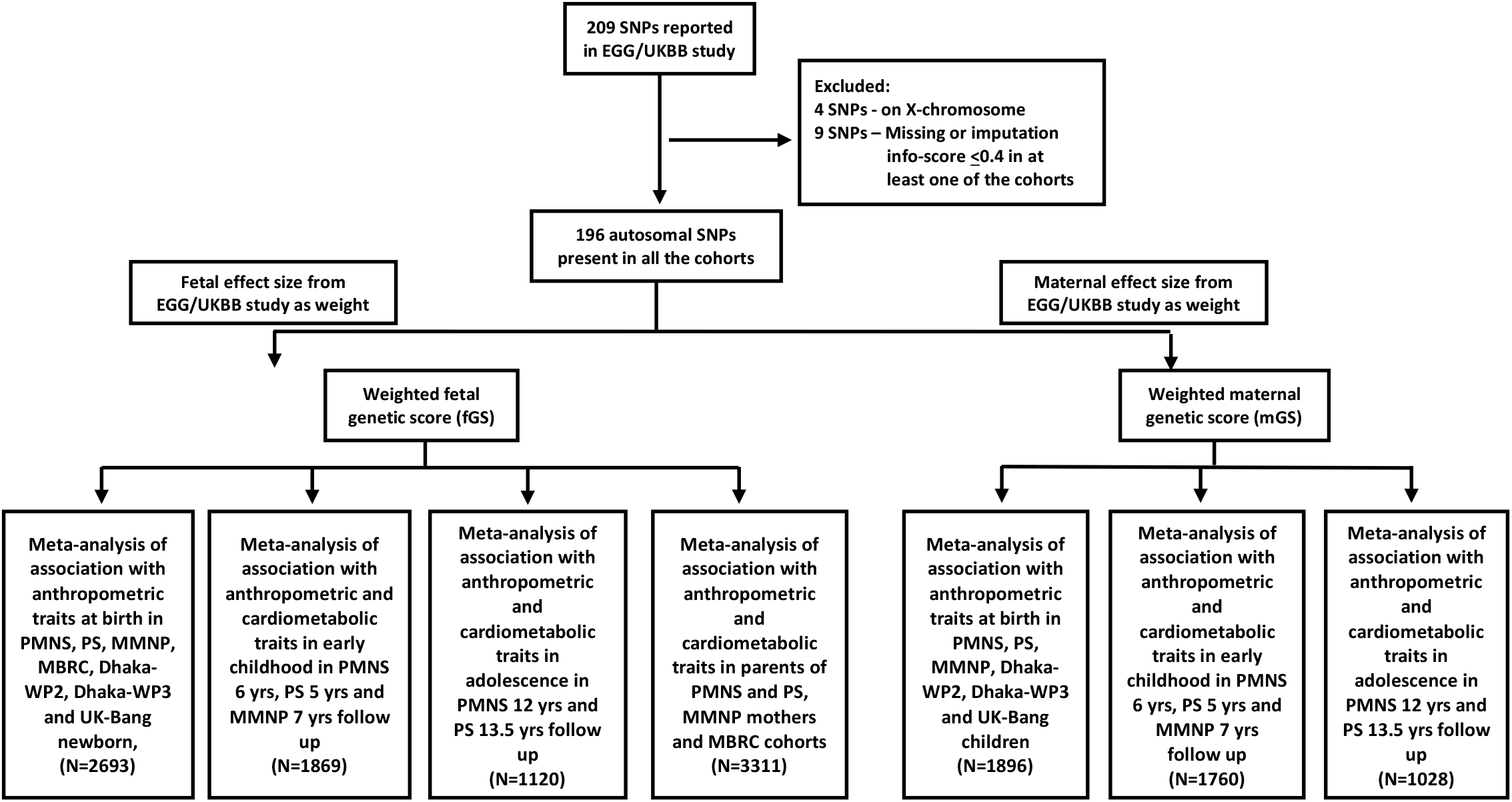
Flow chart showing the overall study design including SNP selection, generation of weighted fetal and maternal genetic scores, association analysis and final meta-analyses at different stages of follow-up. SNP, single nucleotide polymorphism; EGG, Early Growth Genetics Consortium; UKBB, UK Biobank; PMNS, Pune Maternal Nutrition Study; PS, Parthenon Study; MMNP, Mumbai Maternal Nutrition Project; MBRC, Mysore Birth Records Cohort; Dhaka-WP2, Work Package 2 of GIFTS; Dhaka-WP3, Work Package 3 of GIFTS; UK-Bang, London UK Bangladeshi cohort.

### Association of genetic scores with birth weight and other birth measures

The weighted fetal birth weight genetic score calculated from 196 SNPs was associated with birth weight in South Asians (Table 2A). The meta-analysis of the South Asian cohorts showed a 0.013 SD higher birth weight per 1 unit higher fGS, adjusted for the child’s sex and gestational age (P = 9.1×10^−11^) (Figure 2A and Table 2A). This is equivalent to 50.7 g of birth weight per SD unit of fGS (Figure 3A). The strength of association was only partially attenuated after additional adjustment for the mGS (Effect = 0.015 SD, P = 1.1×10^−10^) (Figure 2B and Table 2A). The mGS was also directly associated with offspring birth weight although compared to the fGS association, the effect size was smaller (effect = 0.006 SD, P = 0.003) which is equivalent to 33.7 g of birth weight per SD unit of mGS and adjustment for fGS made little difference (effect = 0.006 SD; P = 0.004) (Figure 2C & 2D, Figure 3B, Table 2B). Similar associations were noted after the exclusion of offspring of GDM women from the analysis (effect = 0.010; P = 5.1×10^−8^ for the fGS and effect = 0.005; P = 0.011 for the mGS) (Supplementary Table S3A and S3B). Analyses of only Indians and only Bangladeshis showed consistent and overlapping effect sizes in the fGS association analysis, but the mGS association with birth weight was largely driven by the Bangladeshi cohorts (Supplementary Table S4A and S4B). The distribution of fGS in the South Asian cohorts was similar to the European cohort, EFSOCH (n = 674). However, the distribution of birth weight was shifted to left in the South Asians. A plot of fGS versus birth weight showed that for each fGS, birth weight was substantially smaller in the South Asians (Figures 4A and 4B). Similar observations were noted for the association of mGS with birth weight (Figures 4C and 4D). The effect sizes of the fGS on birth weight in the South Asian cohorts was comparable to the same in EFSOCH and also with South Asians in the UK Biobank study (UKBB-SAS; n=2732) (P = 0.17; P = 0.23 respectively) (Figure 3A). Similarly, the association between mGS and offspring birth weight in our study was similar to that observed in UK Biobank South Asians (P = 0.93). However, there was a trend for the effect size to be smaller among all the South Asian cohorts in our study combined than in EFSOCH (P = 0.048) (Figure 3B). The fGS was also positively associated with additional birth measures including birth length, ponderal index, head circumference, chest circumference, abdomen circumference, mid-upper arm circumference, triceps and subscapular skinfold thickness; no associations were seen with the mGS (Table 3). Respective adjustments for maternal and fetal genetic score did not substantially change the strength of these associations (Supplementary Table S5).

**Table 2A:**
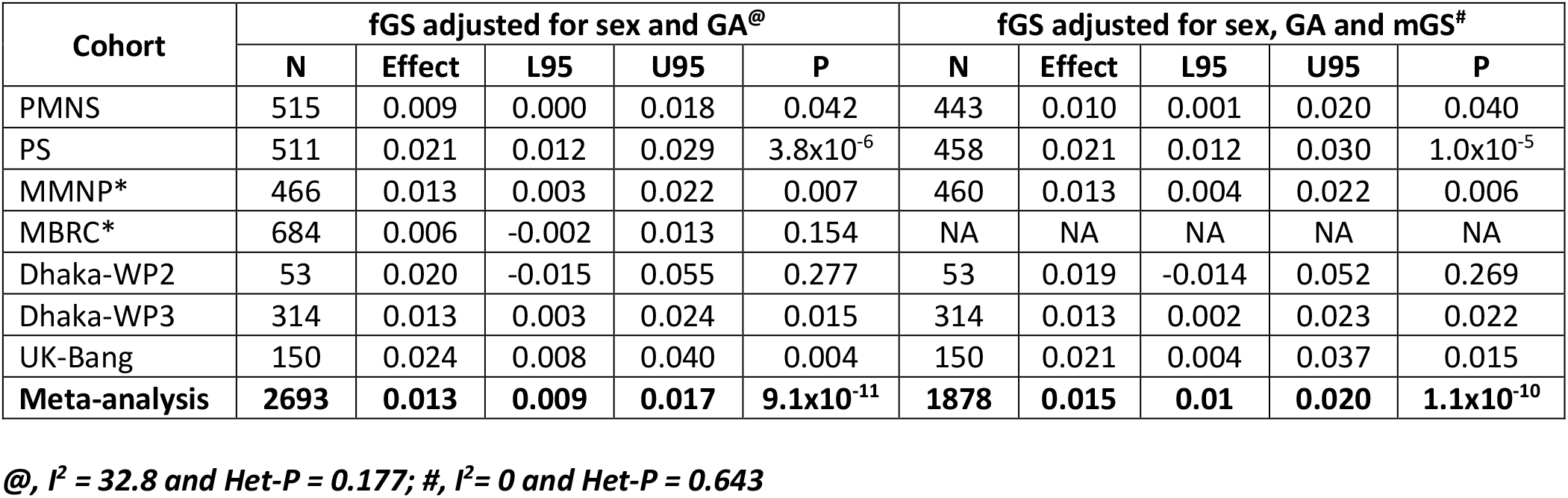
Associations of fetal genetic score (fGS) with own birthweight in South Asian populations.

**Table 2B:** Associations of maternal genetic score with offspring birthweight in South Asian populations.

| Cohort | mGS adjusted for sex and GA<sup>@</sup> |  |  |  |  | mGS adjusted for sex, GA and fGS<sup>#</sup> |  |  |  |  |
| --- | --- | --- | --- | --- | --- | --- | --- | --- | --- | --- |
|  | N | Effect | L95 | U95 | P | N | Effect | L95 | U95 | P |
| PMNS | 461 | 0.000 | -0.008 | 0.008 | 0.976 | 443 | 0.001 | -0.008 | 0.009 | 0.876 |
| PS | 475 | 0.011 | 0.003 | 0.020 | 0.013 | 458 | 0.011 | 0.003 | 0.019 | 0.011 |
| MMNP* | 467 | -0.001 | -0.009 | 0.007 | 0.804 | 460 | 0.000 | -0.009 | 0.008 | 0.957 |
| Dhaka-WP2 | 53 | 0.034 | 0.009 | 0.059 | 0.011 | 53 | 0.034 | 0.009 | 0.059 | 0.011 |
| Dhaka-WP3 | 314 | 0.010 | 0.001 | 0.020 | 0.040 | 314 | 0.009 | 0.000 | 0.019 | 0.060 |
| UK-Bang | 150 | 0.016 | 0.001 | 0.032 | 0.041 | 150 | 0.012 | -0.004 | 0.028 | 0.150 |
| **Meta-analysis** | **1903** | **0.006** | **0.002** | **0.010** | **0.003** | **1878** | **0.006** | **0.002** | **0.010** | **0.004** |

**@, $I^2 = 63.5$ and Het-P = 0.018; #, $I^2 = 53.7$ and Het-P = 0.056**
Association analysis was performed using linear regression with standardized birthweight adjusted for sex and gestational age as the dependent variable for each cohort separately and finally the summary results were meta-analyzed. \*In MMNP, allocation group was additionally adjusted for, and in MBRC only sex was adjusted for, since gestational age data was not available for the majority of the sample. The effect size is in standard deviation units of birthweight per unit change in genetic score. The standard deviation of birthweight in kg in all these cohorts ranged from 0.34 to 0.45 kg. N, number of term babies; GA, gestational age; L95, U95, 95% confidence interval; $I^2$ , heterogeneity; Het-P, P value for heterozygosity; P, P value; fGS, fetal genetic score; mGS, maternal genetic score; GA, gestational age. PMNS, Pune Maternal Nutrition Study; PS, Parthenon Study; MMNP, Mumbai Maternal Nutrition Project; MBRC, Mysore Birth Records Cohort; Dhaka-WP2, Work Package 2 of GIFTS; Dhaka-WP3, Work Package 3 of GIFTS; UK-Bang, London UK Bangladeshi cohort.

**Table 3:** Associations of fetal and maternal genetic scores with other birth measures in South Asian populations.

| Trait | fGS adjusted for sex and gestational age* |  |  |  |  |  |  | mGS adjusted for sex and gestational age* |  |  |  |  |  |  |
| --- | --- | --- | --- | --- | --- | --- | --- | --- | --- | --- | --- | --- | --- | --- |
|  | N | Effect | L95 | U95 | P | I<sup>2</sup> | Het-P | N | Effect | L95 | U95 | P | I<sup>2</sup> | Het-P |
| Birth length (Z) | 2544 | 0.004 | 0.000 | 0.009 | 0.048 | 44.1 | 0.097 | 1820 | 0.003 | -0.002 | 0.008 | 0.153 | 42.5 | 0.122 |
| Ponderal Index (Z) | 2517 | 0.009 | 0.004 | 0.013 | 2.1x10<sup>-4</sup> | 28.3 | 0.213 | 1796 | 0.000 | -0.004 | 0.006 | 0.906 | 14.3 | 0.323 |
| Head circumference (Z) | 2564 | 0.005 | 0.000 | 0.009 | 0.030 | 48.0 | 0.073 | 1844 | 0.002 | -0.002 | 0.007 | 0.425 | 0 | 0.741 |
| Chest circumference (Z) | 1586 | 0.012 | 0.007 | 0.017 | 8.2x10<sup>-6</sup> | 23.1 | 0.273 | 1477 | 0.002 | -0.002 | 0.007 | 0.383 | 3.7 | 0.374 |
| Abdominal circumference (Z) | 1586 | 0.014 | 0.008 | 0.019 | 3.4x10<sup>-7</sup> | 68.5 | 0.023 | 1477 | 0.002 | -0.003 | 0.007 | 0.554 | 62.0 | 0.048 |
| Mid-upper arm circumference (Z) | 1953 | 0.014 | 0.009 | 0.019 | 1.3x10<sup>-7</sup> | 0 | 0.485 | 1844 | 0.005 | 0.000 | 0.010 | 0.045 | 0 | 0.982 |
| Triceps skinfold (Z) | 1564 | 0.013 | 0.007 | 0.018 | 3.6x10<sup>-6</sup> | 44.6 | 0.144 | 1455 | 0.003 | -0.001 | 0.009 | 0.181 | 61.7 | 0.050 |
| Subscapular skinfold (Z) | 1563 | 0.012 | 0.006 | 0.017 | 2.4x10<sup>-5</sup> | 42.3 | 0.158 | 1454 | 0.003 | -0.002 | 0.008 | 0.260 | 25.7 | 0.258 |

Association analysis was performed using linear regression with standardized birth measures adjusted for sex and gestational age as the dependent variables for each cohort independently and finally the summary results were meta-analyzed. The effect size is in standard deviation units of the birth measure per unit change in genetic score. The South Asian populations include PMNS, Pune Maternal Nutrition Study; PS, Parthenon Study; MMNP, Mumbai Maternal Nutrition Project from India; MBRC, Mysore Birth Records Cohort; Dhaka-WP2 of GIFTS; Dhaka-WP3 of GIFTS; UK-Bang, London UK Bangladeshi cohort. \*In MMNP, allocation group was additionally adjusted for, and in MBRC only sex was adjusted for since gestational age data was not available for the majority of the sample. N, number of term babies; L95, U95, 95% confidence interval; I<sup>2</sup>, heterogeneity; Het-P, P value for heterozygosity; P, P value; fGS, fetal genetic score; mGS, maternal genetic score. The N was different for each trait due to missingness of some phenotype data in MBRC, Dhaka-WP2 and Dhaka-WP3.

**Figure 2:**
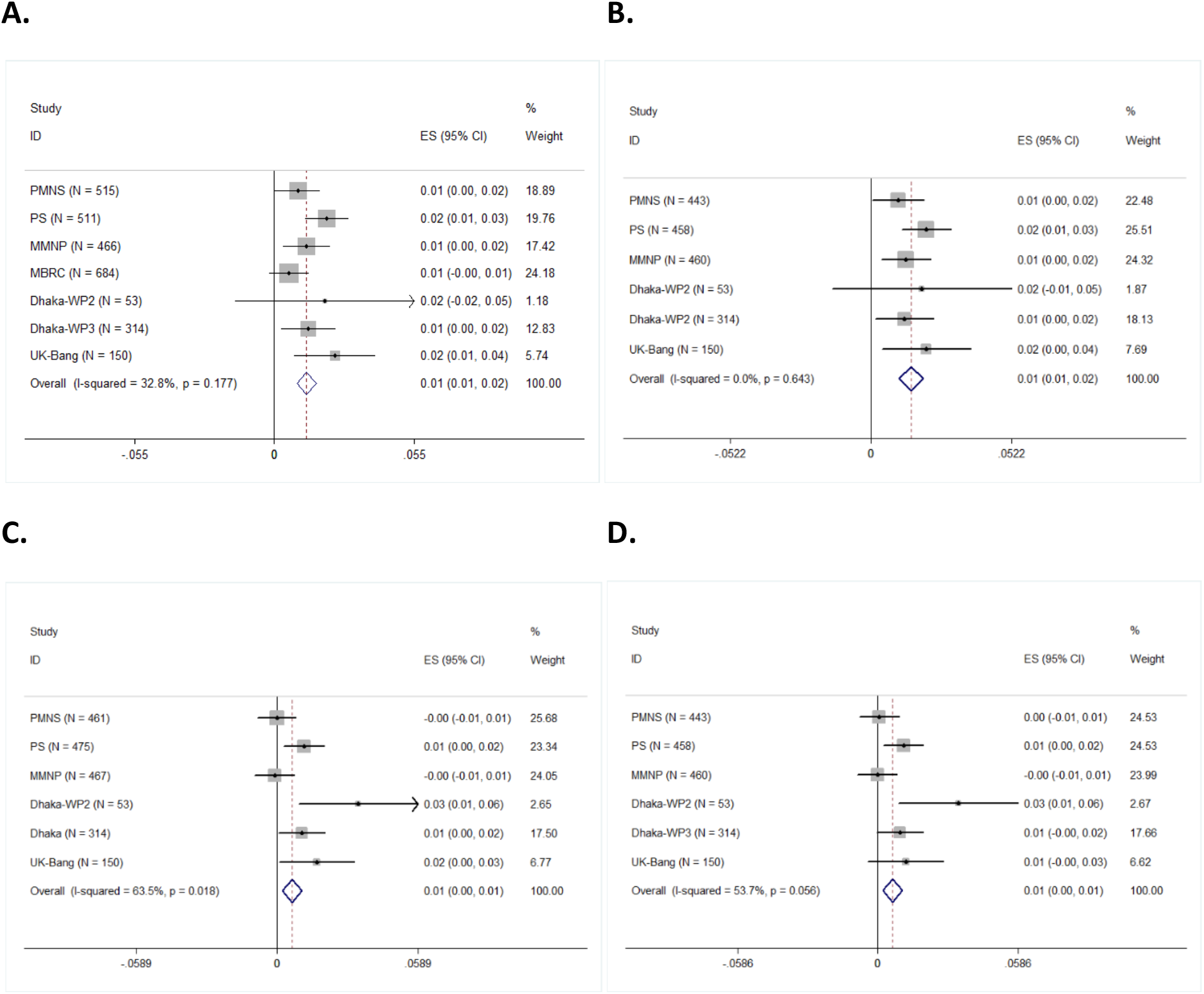
Meta-analysis of associations of weighted genetic scores derived from 196 single nucleotide polymorphisms with birthweight in South Asian populations. (A) fetal genetic score adjusted for sex and gestational age (B) fetal genetic score adjusted for sex, gestational age and maternal genetic score (C) maternal genetic score adjusted for sex of the fetus and gestational age and (D) maternal genetic score adjusted for sex of the fetus, gestational age and fetal genetic score. *In MMNP, allocation group was additionally adjusted for and in MBRC, only sex was adjusted for, since gestational data was not available for the majority of the sample. PMNS, Pune Maternal Nutrition Study; PS, Parthenon Study; MMNP, Mumbai Maternal Nutrition Project; MBRC, Mysore Birth Records Cohort; Dhaka-WP2, Work Package 2 of GIFTS; Dhaka-WP3, Work Package 3 of GIFTS; UK-Bang, London UK Bangladeshi cohort; ES, effect size; CI, confidence interval; I^2^, heterogeneity; p, P-value.

**Figure 3:**
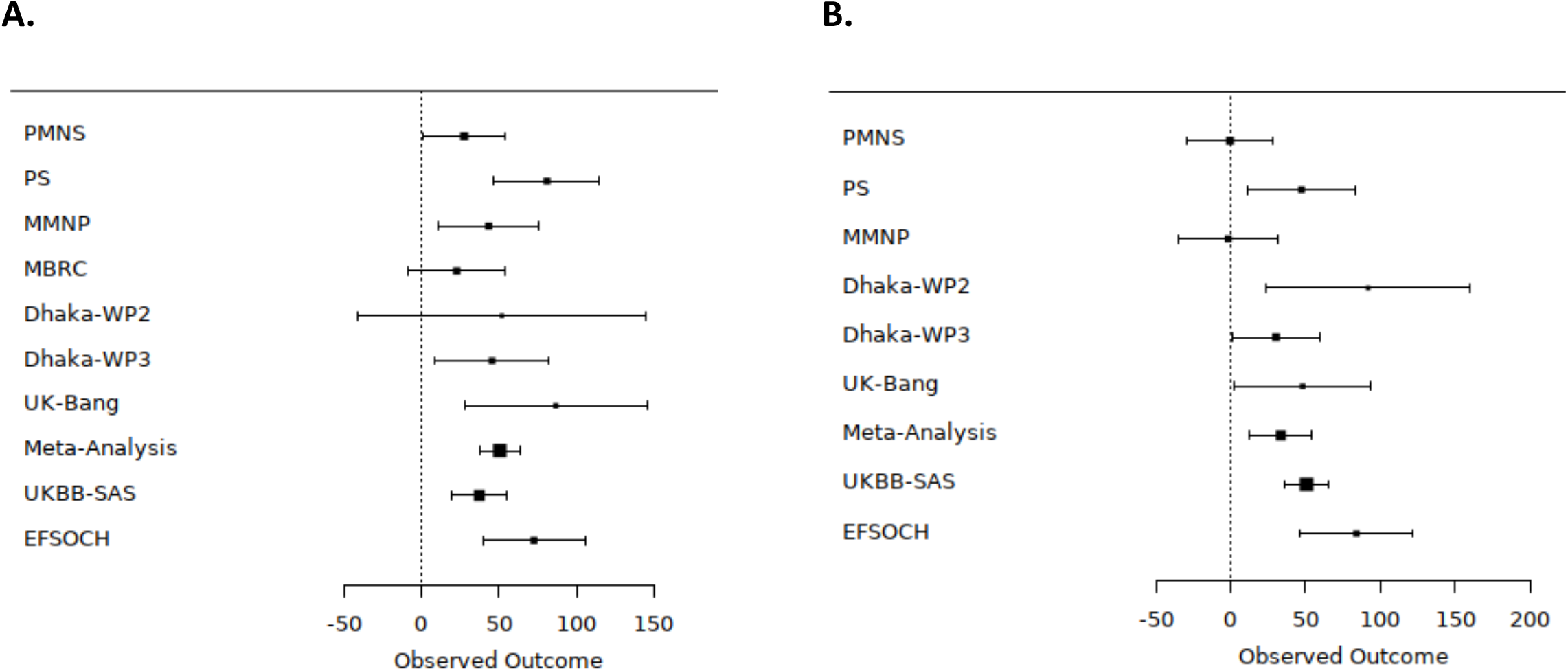
Comparison of the effect of (A) weighted fetal genetic score (B) weighted maternal genetic score, on birthweight in different South Asians cohorts in the present study and the UK Biobank South Asian component (UKBB-SAS) and The Exeter Family Study of Childhood Health (EFSOCH) study, a European study with similar sample size. The X-axis indicates the effect size for birthweight in gram per standard unit of weighted genetic score. PMNS, Pune Maternal Nutrition Study; PS, Parthenon Study; MBRC, Mysore Birth Records Cohort; MMNP, Mumbai Maternal Nutrition Project; Dhaka-WP2, Work Package 2 of GIFTS; Dhaka-WP3, Work Package 3 of GIFTS; UK-Bang, London UK Bangladeshi cohort. Heterogeneity P value for fGS is 0.1777 and for mGS is 0.0046.

**Figure 4:**
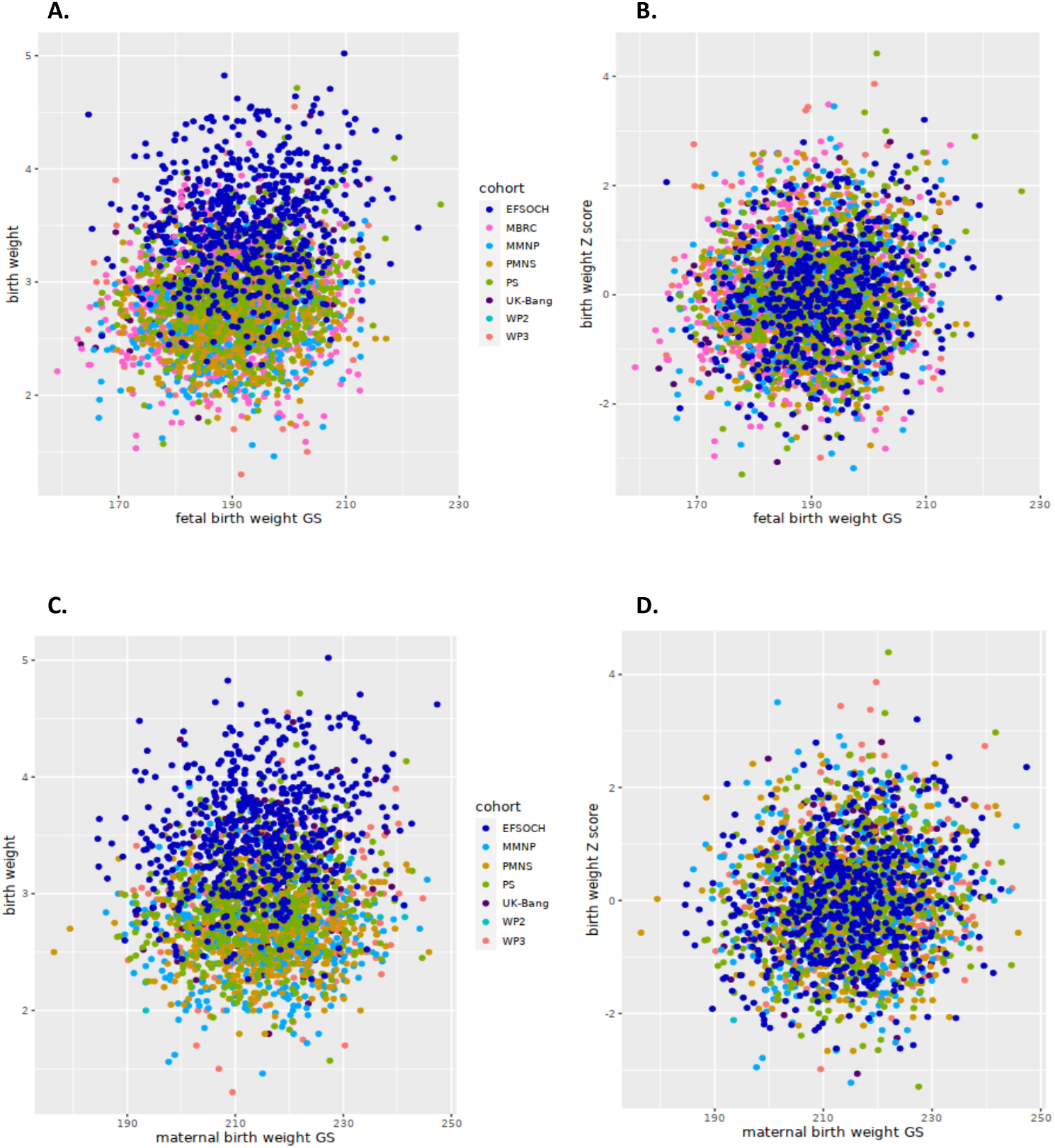
Scatter plot comparing the correlation between birthweight and fetal genetic score (fGS)(A, B)& birthweight and maternal genetic score (mGS) (C, D) in the South Asian cohorts (PMNS, Pune Maternal Nutrition Study; PS, Parthenon Study; MMNP, Mumbai Maternal Nutrition Project; MBRC, Mysore Birth Records Cohort; Dhaka-WP2, Work Package 2 of GIFTS; Dhaka-WP3, Work Package 3 of GIFTS; UK-Bang, London UK Bangladeshi cohort) and an European Cohort (EFSOCH, The Exeter Family Study of Childhood Health). A, indicates absolute birthweight and fGS; B, shows the same between cohort-specific birthweight Z-scores and fGS; C, indicates absolute birthweight and mGS and D, shows the same between cohort-specific birthweight Z-scores and mGS.

### Associations of birth weight and fetal genetic score with anthropometric and cardiometabolic traits in follow-up stages

The associations of birth weight and the fGS with later anthropometric and cardiometabolic traits in early childhood and early adolescence were investigated in the Indian cohorts only, since they had longitudinal follow-up data. Birth weight was strongly positively associated with all anthropometric traits in childhood (5-7 years; P = 3×10^−5^ - 1.9×10^−51^) and adolescence (11-14 years; P = 5.7×10^−6^ – 8.1×10^−27^) (Figure 5; Supplementary Table S6). It also showed strong evidence of a negative association with triglyceride levels in childhood (P=9.8×10^−4^) and weakly in adolescence (P=0.002). We observed a negative association with systolic and diastolic blood pressure and a positive association with fat percentage both in childhood and adolescence but these did not pass the Bonferroni-corrected threshold of P<0.001 (Supplementary Table S6). Similarly, a higher fGS was associated with larger body size in the children (Table 4). We observed a strong positive association of the fGS with waist circumference (effect = 0.01 SD per standard unit, P = 5.7×10^−5^) but the associations with other anthropometric parameters including weight, height, BMI, head circumference and mid-upper arm circumference were weaker (P = 0.017 – 0.001) and did not pass the multiple testing threshold of P<0.001 (Table 4; Figure 6)]. No strong evidence of associations between fGS and anthropometric traits were detected in adolescents. The fGS was not associated with any of the cardiometabolic parameters in children or in adolescents (Table 4). The mGS had no influence on anthropometric and cardiometabolic parameters in children or in adolescents (Supplementary table S7).

**Table 4:** Meta-analysis^#^ of associations of fetal genetic score with anthropometric and cardiometabolic traits in early childhood, adolescence and adults in Indians.

| Traits | Children |  |  |  |  | Adolescents |  |  |  |  | Adults |  |  |  |  |
| --- | --- | --- | --- | --- | --- | --- | --- | --- | --- | --- | --- | --- | --- | --- | --- |
|  | N | Effect | P | I<sup>2</sup> | Het-P | N | Effect | P | I<sup>2</sup> | Het-P | N | Effect | P | I<sup>2</sup> | Het-P |
| Weight (Z) | 1866 | 0.008 | 0.001 | 0 | 0.830 | 1120 | 0.002 | 0.592 | 0 | 0.641 | 3311 | 0.002 | 0.341 | 0 | 0.698 |
| Height (Z) | 1865 | 0.006 | 0.017 | 0 | 0.846 | 1120 | 0.002 | 0.437 | 0 | 0.889 | 3307 | 0.003 | 0.037 | 0 | 0.574 |
| Body mass index (Z) | 1865 | 0.007 | 0.007 | 0 | 0.666 | 1120 | 0.001 | 0.844 | 0 | 0.581 | 3306 | 0.000 | 0.977 | 0 | 0.438 |
| Head circumference (Z) | 1866 | 0.007 | 0.003 | 0 | 0.999 | 1115 | 0.004 | 0.223 | 0 | 0.633 | 3256 | 0.006 | 5.5x10<sup>-4</sup> | 32.2 | 0.194 |
| Waist circumference (Z) | 1864 | 0.010 | 5.5x10<sup>-5</sup> | 0 | 0.463 | 1096 | 0.004 | 0.254 | 0 | 0.918 | 3251 | 0.001 | 0.528 | 13.8 | 0.326 |
| Hip circumference (Z) | NA | NA | NA | NA | NA | NA | NA | NA | NA | NA | 3256 | 0.001 | 0.456 | 0 | 0.680 |
| Waist to hip ratio (Z) | NA | NA | NA | NA | NA | NA | NA | NA | NA | NA | 3247 | 0.001 | 0.603 | 9.8 | 0.353 |
| Mid upper arm circumference (Z) | 1865 | 0.005 | 0.032 | 0 | 0.705 | 1112 | 0.000 | 0.976 | 0 | 0.595 | 3258 | 0.000 | 0.852 | 0 | 0.645 |
| Triceps skinfold (Z) | 1865 | 0.002 | 0.511 | 0 | 0.760 | 1114 | 0.002 | 0.487 | 0 | 0.790 | 3259 | 0.001 | 0.748 | 0 | 0.725 |
| Subscapular skinfold (Z) | 1865 | 0.003 | 0.280 | 52.2 | 0.123 | 1113 | 0.002 | 0.603 | 0 | 0.825 | 3238 | -0.001 | 0.673 | 0 | 0.926 |
| Fat percentage (Z) | 1860 | 0.003 | 0.254 | 50.3 | 0.133 | 1085 | 0.002 | 0.475 | 45.8 | 0.174 | NA | NA | NA | NA | NA |
| Systolic blood pressure (Z)* | 1847 | -0.002 | 0.411 | 0 | 0.410 | 1102 | -0.005 | 0.112 | 88.6 | 0.003 | 3081 | 0.000 | 0.801 | 0 | 0.454 |
| Diastolic blood pressure (Z)* | 1848 | 0.000 | 0.989 | 0 | 0.765 | 1102 | 0.000 | 0.904 | 92.4 | 0.000 | 3082 | 0.000 | 0.922 | 0 | 0.467 |
| Fasting glucose (Z)* | 1840 | -0.002 | 0.483 | 0 | 0.497 | 1110 | 0.000 | 0.908 | 92.8 | 0.000 | 2601 | -0.006 | 9.3x10<sup>-4</sup> | 30.5 | 0.218 |
| 120 minutes glucose (Z)* | 1809 | 0.002 | 0.321 | 0 | 0.434 | NA | NA | NA | NA | NA | 1320 | 0.000 | 0.905 | 0 | 0.707 |
| Fasting insulin (Z)* | 1831 | 0.002 | 0.369 | 18.9 | 0.291 | 1111 | 0.002 | 0.463 | 47.7 | 0.167 | 2596 | -0.002 | 0.359 | 0 | 0.823 |
| HOMA-IR (Z)* | 1756 | 0.002 | 0.401 | 0 | 0.997 | 1110 | 0.002 | 0.407 | 74.4 | 0.048 | 2432 | -0.005 | 0.022 | 0 | 0.802 |
| Total cholesterol (Z)* | 1838 | -0.005 | 0.050 | 50.7 | 0.131 | 1111 | 0.004 | 0.224 | 0 | 0.488 | 2601 | -0.003 | 0.118 | 0 | 0.968 |
| LDL-cholesterol (Z)* | 1847 | -0.003 | 0.280 | 52.9 | 0.119 | 1111 | 0.006 | 0.070 | 0 | 0.676 | 2600 | -0.001 | 0.594 | 0 | 0.957 |
| HDL cholesterol (Z)* | 1849 | -0.005 | 0.059 | 0 | 0.513 | 1111 | 0.002 | 0.632 | 0 | 0.631 | 2584 | 0.000 | 0.867 | 0 | 0.809 |
| Triglycerides (Z)* | 1838 | -0.001 | 0.666 | 0 | 0.668 | 1111 | -0.002 | 0.440 | 37.8 | 0.205 | 2601 | -0.006 | 0.002 | 0 | 0.673 |

Association analysis was performed using linear regression with standardized log10 transformed traits as the dependent variable for each cohort independently and finally the summary results were meta-analyzed. Age and sex were included as covariates in the regression model for all traits; BMI was additionally included as a covariate for analysis of traits marked with an asterisk (\*). Allocation group was additionally adjusted for in MMNP. #, Meta-analysis for children included those from Pune Maternal Nutrition Study at 6 yrs, Parthenon Study at 5 yrs, and Mumbai Maternal Nutrition Project at 7 yrs of age; for adolescents from Pune Maternal Nutrition Study at 12 yrs and Parthenon Study at 13.5 yrs; and for adults from parents from Pune Maternal Nutrition Study and Parthenon Study, mothers from Mumbai Maternal Nutrition Project, and individuals from Mysore Birth Records Cohort; P, P value; I<sup>2</sup>, heterogeneity; Het-P, P value for heterozygosity; SNP, single nucleotide polymorphism; HOMA-IR, homeostasis model assessment of insulin resistance, LDL, low density lipoprotein; HDL, high density lipoprotein, NA, not available. Those passing the Bonferroni corrected P<sub>≤</sub>0.001 were considered as statistically significant.

**Figure 5:**
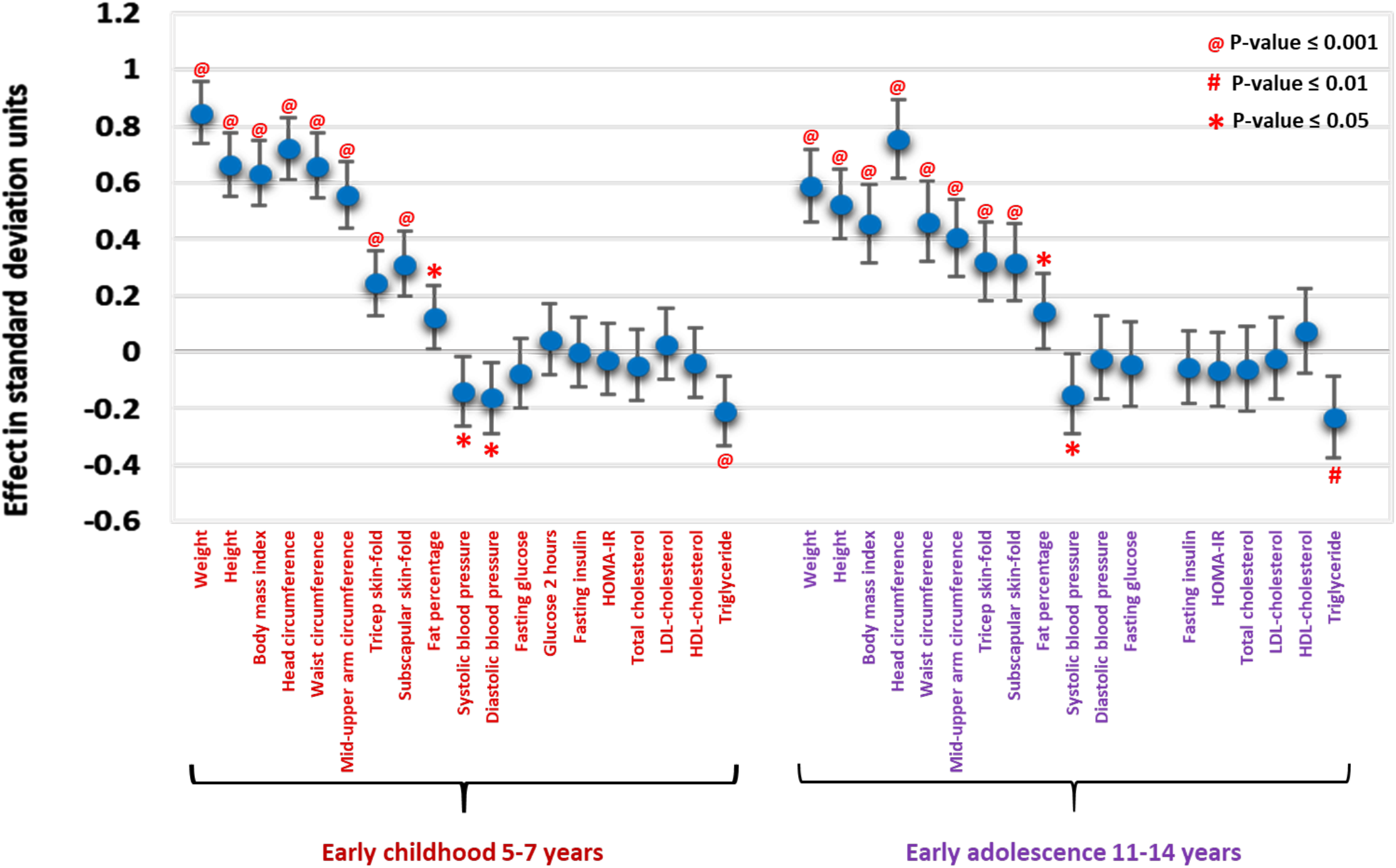
Associations of birthweight with a range of anthropometric and cardiometabolic traits during early childhood and early adolescence in the Indian cohorts. The X-axis shows anthropometric and cardiometabolic traits at different stages of follow-up and the Y-axis indicates the effect size in standard deviation units. HOMA-IR, Homeostasis Model Assessment of Insulin Resistance; LDL, low density lipoprotein; HDL, high density lipoprotein. Please see figure 1 for details of cohorts used at different life stages.

**Figure 6:**
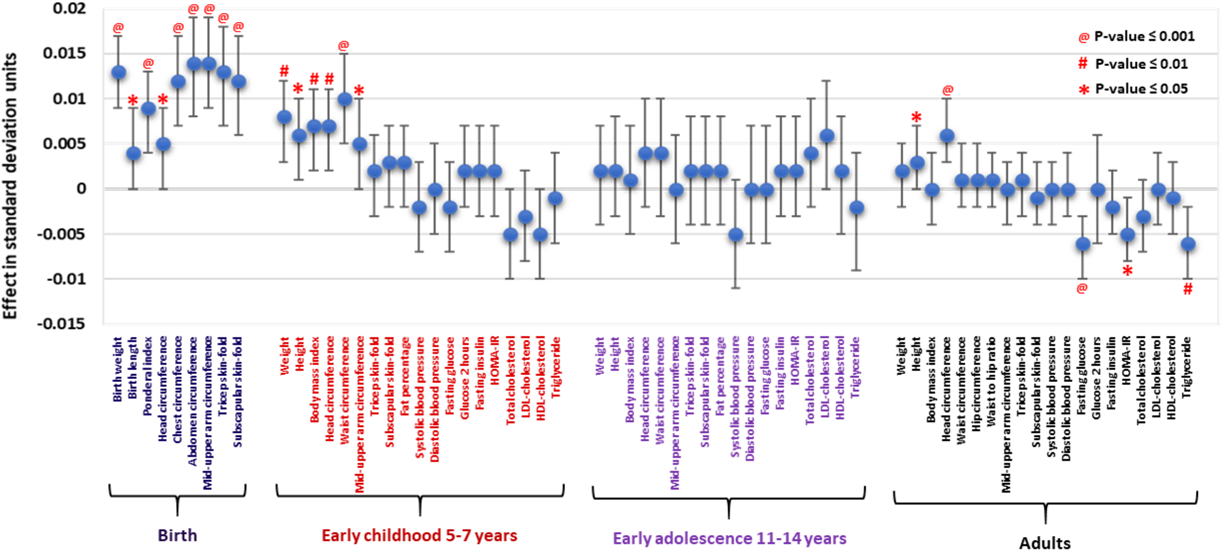
Associations of fetal genetic score with various birth measures and a range of anthropometric and cardio-metabolic traits during early childhood, adolescence and adults in the Indian cohorts. The X-axis shows anthropometric and cardio-metabolic traits at different stages of follow-up and the Y-axis indicates the effect size in standard deviation units. HOMA-IR, Homeostasis Model Assessment of Insulin Resistance; LDL, low density lipoprotein; HDL, high density lipoprotein; EGG, Early Growth Genetics Consortium; UKBB, UK Biobank. Please see figure 1 for details of cohorts used at different life stages.

Using data on parents of children in the PMNS & PS, men and women in the MBRC and mothers in the MMNP cohort, we investigated the influence of fGS on anthropometric and cardiometabolic traits in adults (Figure 6, Table 4). The fGS showed a strong positive association with head circumference (effect = 0.006; P = 5.5×10^−4^) and a weak positive association with adult height (effect = 0.002; P = 0.037) (Table 4; Figure 6). It was also negatively associated with fasting glucose (effect = −0.006; P = 9.3×10^−4^) and showed a weak negative association with HOMA-IR and triglycerides (P = 0.022 and 2.0×10^−3^ respectively). The direction of associations was the same as the genome-wide genetic correlations reported in Warrington NM et al. 2019 (P ranging from 0.002 - 5.5×10^−4^) (Figure 6; Table 4). No evidence of associations was noted with the other anthropometric and cardiometabolic traits in adults (P>0.05) (Table 4).

## DISCUSSION

In this study which included several Indian and Bangladeshi cohorts from both the UK and Indian subcontinent, we investigated whether the genetic variants identified in a GWAS of birth weight in Europeans also influence birth size in South Asians. We further investigated whether the same genetic variants (either fetal variants that directly influence birth weight, or those in the mother that act indirectly via the intrauterine environment) were associated with serially measured anthropometric and cardiometabolic parameters during early childhood and adolescence. We observed strong positive associations of fetal genetic score with birth weight and other birth measurements in these South Asian populations despite the large variation in maternal BMI and fetal birth weight. Birth weight was positively related to all body measurements during childhood and so was the fetal genetic score. While birth weight predicted body size in both children and adolescents, fetal genetic score did so in children but not adolescents. We also noted a strong association of birth weight with plasma triglyceride levels both in children and adolescents, but fetal genetic score was not related to any of the child/adolescent cardiometabolic outcomes. Maternal genetic score was weakly positively related to birth weight and was unrelated to body size and cardiometabolic traits in both children and adolescents. Fetal genetic score was inversely related to fasting plasma glucose and triglycerides in the adults, however maternal genetic scores were not available for the adults and birth weight was not available for a sufficient number of the adults to examine birth weight effects on body size and cardiometabolic parameters. Our study thus replicates the strong association of fetal genetic score and weak association of maternal genetic score (developed from European mother-child dyads) with birth weight and other birth measures in a non-European population. We also observe that the genetic constitution of the fetus at specific variants influences body size and the data from the adults suggests that it contributes to future cardiometabolic risk in Indians. Follow up studies on a larger sample size will be required to answer our second research question (is the birth weight – cardiometabolic risk association explained by shared genetic variants) with confidence.

Most genetic studies associating early life parameters with future risk of cardiometabolic disorders have been conducted in Europeans. As far as we are aware, this is first such analysis in South Asians. We found similar associations of fetal genetic scores generated in the Europeans with birth size in a consortium of 7 birth cohorts comprising Indian and Bangladeshi mother-child pairs. This was despite a wide variability in birth weight and maternal BMI within the South Asian cohorts and significant differences in the effect allele frequency of many of the birth weight associated variants between the EGG/UKBB and the South Asian subjects. It was interesting to note that despite similar fetal genetic risk scores, there was a large difference in birth weight between South Asian and European populations. There was an increase of 50.7 g of birth weight per SD of the fetal genetic score which is consistent with that observed in the UKBB-SAS and marginally smaller than in EFSOCH, examples of South Asian and European cohorts respectively. The significant association of fetal genetic score with body size at birth persisted even after adjustment for maternal genetic score, indicating that the genetic effect is not influenced by aspects of the intrauterine environment predicted by the genetic variants used in this study. This is further supported by a similar strength of association after exclusion of children born to GDM mothers; which suggests that the fetal genetic effects are independent of maternal diabetes status during pregnancy. Consistent with the observations in Europeans, birth weight was only weakly influenced by the maternal genetic score and this was unaffected by the fetus’s own genotype suggesting that the maternal genetic effect on birth weight was mediated through intrauterine environment. Thus, birth weight (body size) is an outcome of the baby’s genetic constitution and an influence of the intrauterine environment, partly determined by the mother’s genotype. However, with the exception of a small number of variants that are known to influence fasting glucose levels, it is largely unknown which intrauterine exposures are influenced by which genetic variants used in the study, making it difficult to dissect their relative role. It was interesting to note that the influence of the maternal genetic score on birth weight varied considerably amongst the cohorts investigated in this study (P=0.018). The cause of this heterogeneity in effect estimates could be driven by ethnicity, maternal BMI, height and nutritional status, socio-economic status, and GDM status; this needs further investigation.

Genome-wide studies have established a robust association between birth weight genetic score and later cardiometabolic risk including glycaemic and lipid parameters in Europeans^12, 14^. An important feature of our study is that we have been able to independently compare associations of birth weight and birth weight-associated genetic variations on later anthropometric and cardiometabolic traits. Birth weight showed a strong positive association with body composition, and a directionally inverse association with blood triglyceride concentrations in both childhood and adolescence. Despite the fact that fetal genetic score explains only 4% of the variance in birth weight in European Individuals^14^, it showed a positive association with body size in early childhood, height and head circumference in adults. Effect estimates of fetal genetic score with other anthropometric traits was directionally consistent with the direct effect of birth weight; a lack of strong association may be due to a relatively smaller sample size and the smaller effect size compared to the birth weight itself. Absence of association between fetal genetic score and any of the traits during adolescence is consistent with findings from larger studies that have found little evidence of influence of fetal birth weight variants on BMI beyond early childhood^19^. Previous studies, in contrast, have demonstrated positive genetic correlations between birth weight and both childhood and adulthood height^12, 14^. Maternal genetic score is also associated with birth weight but its contribution is even smaller than direct effects of fetal genetic score and has no influence on body size or cardiometabolic risk at any stages. As we do not have estimates of independent maternal and fetal genetic associations with these traits, the interpretation of the associations is not completely clear. However, despite similar genetic scores as in Europeans, South Asians are substantially smaller indicating environmental constraint. The fact that the fetus’s genotype at birth weight associated genetic variants also influenced plasma glucose and triglycerides in adulthood is consistent with the fetal insulin hypothesis, which proposes that birth weight and later cardiometabolic risk are two effects of the same genotype^20^. Our findings need to be replicated in larger independent studies of South Asian subjects. Further understanding of the link between birth weight and future cardiometabolic risk will be possible as we understand the exact role of each genetic variant, whether it operates directly or indirectly through its effects on intrauterine environment.

Our study has several strengths and a few limitations. This is the first study exploring the influence of fetal and/or maternal genotype in determining birth size and their role in future cardiometabolic risk in South Asians. We have combined several cohorts from India (including both Indo-European and Dravidian ethnicities) as well as from Bangladesh, hence the observations can be considered representative of South Asians. The biggest strength of the study is availability of mother-child pairs and serial anthropometric and cardiometabolic traits in early childhood and adolescence and hence the conclusions drawn from these prospective cohorts are robust. A limitation of the study is a relatively small sample size, which may not be adequately powered to detect many associations reported in Europeans. Another limitation is the lack of availability of adult phenotype data on these cohorts, as they are just now reaching adulthood. We have been able to partly circumvent this lack of adult data by using the genotype and phenotype data from parents of the children in the Indian cohorts, but we lacked birth size and maternal genotype data for these individuals, making the analysis less complete than for the children.

The observations made in this study are important because the sub-continent is facing the twin burden of poor fetal health and an emerging increase of type 2 diabetes and cardiovascular disorders^21, 22, 23^. This has been linked to unique phenotypic features, environmental exposures, and a different genetic makeup of South Asians compared to Europeans^16, 17, 18, 24, 25^. However, from this study it seems that the genetic contribution to birth size is largely similar and other factors may be responsible for the differences in the thin fat phenotype of South Asians which predisposes them to a higher risk of diabetes and related disorders compared to Caucasians. The validation of genetic associations in different ethnic populations suggested that there may be common pathways affecting fetal development which can be influenced by different environmental exposures.

To conclude, we have replicated the associations of genetic loci reported in Europeans with size at birth in participants of South Asian ancestry. However, fetal genetic score is known to explain less than 5% of the variability in birth weight in Europeans. Furthermore, despite similar fetal genetic scores as in Europeans, South Asians in this study have considerably lower birth weight, indicating strong environmental constraints on fetal growth. These genetic loci also influenced early childhood body size and in adults were associated with fasting glucose and triglycerides levels, suggesting that common genetic variants could explain part of the association between size at birth and adult metabolic syndrome. This supports the “Fetal insulin hypothesis” but also highlights an important interaction with environment^15, 20^. However, we did not find associations between fetal genetic scores and cardiometabolic traits in the children and adolescents. Further understanding of the link between birth weight and future cardiometabolic risk will require larger studies and will become possible as we understand the precise role of each genetic variant and whether it operates directly or indirectly through its effects on intrauterine environment. Further, associations between fetal genotype and birth weight were consistent across all studies but the association with maternal genotype showed evidence of heterogeneity between studies which may be related to differences in maternal size, glycemia and socio-economic status rather than genetic scores.

## METHODS

### Study participants

The participants in this study were mother-child pairs from different prospective birth cohort studies from India, Bangladesh and UK. The Indian cohorts comprise the Pune Maternal Nutrition Study (PMNS), Parthenon Study (PS), Mumbai Maternal Nutritional Project (MMNP) and Mysore Birth Records Cohort (MBRC). The individuals from PMNS and MMNP have Indo-European ancestry, and those from the PS and MBRC have Dravidian ancestry, the two major ethnic populations in the Indian sub-continent^24, 25^. Informed consent was obtained from all participants following the guidelines of Indian Council of Medical Research, Govt. of India, New Delhi. The Bangladeshi cohorts were from a sub-study of a prospective multi-center European Union FP7 project GIFTS (**G**enomic and l**I**festyle predictors of **F**etal ou**T**come relevant to diabetes and obesity and their relevance to prevention strategies in **S**outh Asian people) consisting of work package (WP2), work package (WP3) and London UK Bangladeshi cohort (UK-Bang).

### Pune Maternal Nutrition Study (PMNS)

The PMNS cohort, based in six rural villages near Pune in Western India, was established in 1993 to examine the relationship of maternal health and nutrition during pregnancy to fetal growth and development, and future cardiometabolic risk^26^. Women were recruited pre-conceptionally. A 75gm oral glucose tolerance test was carried out at 28 weeks’ gestation in pregnancy and GDM was diagnosed based on revised WHO 1999 guidelines. Gestational age was based on last menstrual period dates (recorded every 3 months during the pre-conception period) unless it differed from early (<20 weeks’ gestation) ultrasound scan dating by 2 weeks or more, in which case the latter was used. Detailed new born anthropometry was carried out by trained research staff within 72 hours of birth. Multiple follow-up studies have been conducted starting from pre-pregnancy, during pregnancy, at birth, early childhood, adolescence and young adulthood and detailed anthropometric and biochemical data have been collected. At 6 years of age, we measured anthropometry, resting systolic and diastolic blood pressure, plasma glucose and insulin (fasting and after an oral glucose load) and fasting lipids (triglycerides and LDL- and HDL-cholesterol). At 12 years, detailed anthropometry, and measurements of blood pressure, fasting glucose, insulin and lipids were repeated. At both time points, the same measurements were carried out in both parents. We have used these data in the current study.

### Parthenon Study (PS)

The Parthenon study (PS) was established in 1997-98 in Mysore, South India, to examine the long-term effects of maternal glucose tolerance and nutritional status during pregnancy on cardiovascular risk factors and cognition in the offspring^27^. Women (<32 weeks’ gestation) were recruited in the antenatal clinic of the Holdsworth Memorial Hospital, Mysore. Gestational age was assessed using last menstrual period dates collected at recruitment. A 100gm oral glucose tolerance test was carried out at 28-32 weeks’ gestation and GDM was diagnosed based on Carpenter and Coustan criteria^28, 29^. Detailed newborn anthropometry was carried out by trained research staff within 72 hours of birth. At 5 and 13.5 years of age, we measured anthropometry, resting systolic and diastolic blood pressure, plasma fasting glucose and insulin) and fasting lipids (triglycerides and LDL- and HDL-cholesterol). At 5 years, the same measurements were carried out in their mothers and only fasting glucose and insulin in the fathers. These data were used in this study.

### Mumbai Maternal Nutritional Project (MMNP)

The Mumbai Maternal Nutrition Project was a randomised controlled trial, set up in 2006 among women living in slums in the city of Mumbai, Western India with the objective to test whether improving women’s dietary micronutrient quality before and during conception improves birth weight and other related outcomes^29^. Women were recruited before conception. As in the PMNS, gestational age was assessed using a combination of last menstrual period dates (which were collected monthly during the pre-conceptional period) and ultrasound scans conducted before 20 weeks’ gestation. A 75g oral glucose tolerance test was carried out at 28-32 weeks’ gestation and GDM was diagnosed based on revised WHO 1999 guidelines. Trained research staff carried out newborn anthropometry within 10 days of birth. In the current study, we have used the child phenotype data at birth (anthropometry) and in early childhood (5-7-year follow-up), when detailed anthropometry, systolic and diastolic blood pressure, fasting and post-load glucose and insulin, and fasting LDL- and HDL-cholesterol and triglycerides were measured^30^. Maternal anthropometry, blood pressure and fasting plasma glucose and insulin concentrations were also measured at this follow-up.

### Mysore Birth Records Cohort (MBRC)

The MBRC is a retrospective birth cohort of urban men and women born at the CSI Holdsworth Memorial Hospital during 1934-55^31^. They were recruited for the first time as adults in 1993-95 and cardiometabolic risk factors were measured (mean age 47 years)^7^. Birth weight, length and head circumference were obtained from their mothers’ obstetric records. We have included the anthropometric data at birth and cardiometabolic parameters measured between 40 and 70 years. Gestational age was missing in the majority of subjects and gestational diabetes status was not available. Since maternal DNA samples were not available, the analyses were restricted to the direct association of fetal genetic score and their birth measures and later life outcomes.

### GIFTS Dhaka Bangladeshi cohorts (WP2 and WP3)

WP2 samples were collected between 2011 and 2012 in Dhaka, Bangladesh from women attending the Maternal and Child Health Training Institute (MCHTI), a tertiary Government hospital for antenatal care and registration in Dhaka. Primigravid pregnant women who were in the first trimester of their pregnancy (≤14 week gestation), with a singleton pregnancy conceived naturally and who were willing to participate in the study were included in an observational study during pregnancy and immediately post-partum after written consent^32^. GDM was diagnosed based on revised WHO 1999 guidelines. Women with a prior history of type 2 diabetes, or gestational diabetes or pregnancy induced hypertension were excluded. The aim of WP2 was to establish the methods and feasibility of recruitment and follow-up for an interventional study (WP3). WP3 samples were collected between 2014 and 2015 in Dhaka, Bangladesh from pregnant women attending MCHTI who consented to an open-label micro-nutrient supplement trial of vitamin D and vitamin B12 supplementation^33^. All consenting women eligible under the WP2 criteria were included in the study and samples were collected from mother and baby under the same sampling frame as WP2. Women who were diagnosed later in pregnancy with GDM remained in the study.

### London UK Bangladeshi cohort (UK-Bang)

The cohort was set up between 2012-2015 as an exploratory observational study of gestational diabetes and its consequences on offspring. Pregnant women of Bangladeshi origin were recruited from the Royal London Hospital antenatal clinics at 28 weeks gestation at the time of 75 gm OGTT. GDM was diagnosed based on Revised WHO, 1999 guidelines. Women were recruited during routine antenatal care and enriched for the presence of GDM. Women with multiple pregnancies, pre-existing or overt type 1 or type 2 diabetes were excluded. Gestational age was based on ultrasound scan dating. Detailed new born anthropometry was carried out by trained research staff within 72 hours of birth.

### The Exeter Family Study of Childhood Health (EFSOCH)

EFSOCH is a prospective study of children born between 2000 and 2004, and their parents, from a geographically defined region of Exeter, UK. All women gave informed consent and ethical approval was obtained from the local review committee. Details of study protocol, including measurement of birth weight, are described in Knight et al^34^. Maternal and paternal DNA samples were extracted from parental blood samples obtained at the study visit (when the women were 28 weeks pregnant), and offspring DNA was obtained from cord blood at birth. Genotyping and imputation of EFSOCH samples has been described previously^35^.

### UK Bio Bank South Asian Subjects (UKBB-SAS)

The UK Biobank phenotype preparation has been described in detail elsewhere^14^ (Warrington et al 2019). Briefly, a total of 280,315 participants reported their own birth weight in kilograms and 216,839 women reported the birth weight of their first child on at least one assessment centre visit. Multiple birth were excluded where reported. In the absence of gestational data, participants with birth weight values <2.5kg or >4.5kg were considered pre-term births and excluded. In addition to the genotype quality control metrics performed centrally by the UK Biobank, we defined a subset of “South Asian” ancestry samples^36^. To do this, we generated ancestry informative principal components (PCs) in the 1000 genomes samples. The UK Biobank samples were then projected into this PC space using the SNP loadings obtained from the principal components analysis using the 1000 genomes samples. The UK Biobank participants’ ancestry was classified using K-means clustering centred on the three main 1000 genomes populations (European, African, and South Asian). Those clustering with the South Asian cluster were classified as having South Asian ancestry.

### Genotyping and SNP selection

Genotyping was performed using different genome-wide SNP array chips. The parents and children of PMNS and mothers and children of PS were genotyped using Illumina CoreExome-24 Array. The Affymetrix SNP 6.0 chip was used for a few of the PMNS parents. Men and women in MBRC, fathers in the PS and mother-child pairs in the MMNP, WP2, WP3 and UK-Bang were genotyped using the Illumina Global Screening Array. Genome-wide imputation for each cohort was conducted separately. The 1000 Genome Phase 3 was used as an imputation reference panel for imputation of PMNS, PS, MMNP and MBRC data^37^. For the Bangladeshi cohorts, imputation was performed using the Haplotype Reference Consortium (HRC) as the imputation reference panel^38^. The imputation quality score was set at info score ≥ 0.4. Out of the 209 SNPs reported in the recent GWAS of birth weight in the EGG/UKBB, 196 autosomal SNPs were present and passed QC in all the cohorts^14^.

### Genetic score calculations

The 196 autosomal SNPs were used for generating a weighted genetic birth weight score. There were two types of genetic score, the fetal genetic score (fGS) and maternal genetic score (mGS). The SNP weights for generating the genetic scores were taken from the Structural Equation Model (SEM) adjusted effect estimates of the fetal and maternal effects respectively from the recent GWAS of birth weight from the EGG/UKBB consortium^14^. The weighted genetic score was calculated using the following formula: 

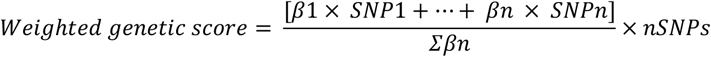

Where β_n_ is the effect size or weight of SNP_n_ taken from the EGG/UKBB birth weight GWAS, nSNPs is the total number of SNP available (n=196), and Σβ_n_ is the sum total weight of all 196 SNPs.

### Statistical analysis

Infants born at less than 37 completed weeks of gestation were excluded. Birth weight and other birth measures were transformed to standardized Z-scores (Z-score = (value – mean)/standard deviation). Association analysis was performed by linear regression, using Z-scores as the dependent variables and weighted genetic score as the independent (predictor) variable, adjusted for the child’s sex and gestational age. The analysis was conducted independently for each cohort and final results were combined by fixed effect inverse variance weighted meta-analysis using the metan command in STATA. The linear regression models were as follows.

#### For the fetal analysis

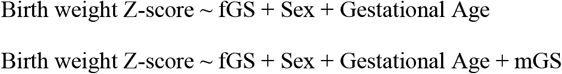

#### For the maternal analysis

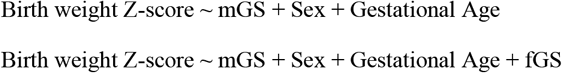

Association analysis of the anthropometric and cardiometabolic phenotype data at follow-up were performed by linear regression, using log10 transformed standardized Z-scores as the dependent variables and weighted genetic score as an independent variable, adjusted for sex and age. BMI was included as an additional covariate for the cardiometabolic traits. The linear regression models were as follows:

#### For the anthropometric traits

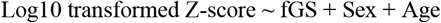

#### For the cardiometabolic traits

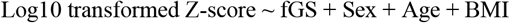

As earlier, the analyses at different follow-up stages were conducted independently for each cohort and fixed effect inverse variance weighted meta-analysis using the metan command in STATA was used to combine the final results. A total of 57 tests in the three stages (childhood, early adolescence and adulthood) were conducted and the significance level was set at P < 0.001 (α < 0.05/57 tests) to allow for multiple testing. Imputed genotype data from fathers and mothers of children in the PMNS and PS, mothers of children in MMNP, and men and women in MBRC were also utilized for investigating the effect of the genetic risk scores on adult anthropometric and cardiometabolic phenotypes.

## Supporting information

STROBE Checklist EQUATOR

## Data Availability

Weights for SNPs used in the genetic score were taken from Warrington et al. 2019 (Nat Genet 51, 804-814 (2019)). We used individual participant data for the association analysis from the PMNS, MMNP, PS, GIFTS, UKB and EFSOCH cohorts. Association summary statistics of genetic score with birth weight of all the cohorts are provided in the manuscript.
Requests for access to the original dataset should be made in writing in the first instance
for PMNS, PS, MMNP and MBRC to the Indian cohorts team leader Giriraj R Chandak at
For the original EFSOCH dataset to the EFSOCH data team via the Exeter Clinical Research Facility
For GIFTS (WP2 & WP3) and UK Bang, to the GIFTS investigator team via Graham A Hitman
Full information on how to access the UK-biobank data can be found at - https://www.ukbiobank.ac.uk/using-the-resource/ [ukbiobank.ac.uk]

## Acknowledgements

We sincerely acknowledge the unflinching support of individuals who voluntarily participated in the study and continued to attend regular follow-ups. S.S.N. and A.D. are grateful to the Council of Scientific and Industrial Research (CSIR Mission Mode Project; HCP0008) and A.S. to the Indian Council for Medical Research (ICMR-CAR; GAP0504), Govt of India for their fellowship. We acknowledge the Genetic Laboratory, Erasmus MC, University Medical Centre Rotterdam for the help with genotyping of the Bangladeshi cohorts. Thanks are also due to the UK Medical Research Council Clinical Research Training Fellowship (No. G0800441) to S.F.

## Authors’ contributions

G.R.C., C.S.Y., G.A.H., C.H.D.F., S.F. and R.M.F. conceived and contributed to the study design; collated and interpreted overall results from various cohorts in the study. G.V.K., K.K., S.A.S., R.D.P., M.K., C.D.G., C.S.Y. and C.H.D.F. are coordinators for various Indian cohorts and played important role in the follow-up and acquisition of phenotype data at different stages. G.R.C. supervised the overall Indian cohort studies. S.F., G.A.H. are the lead supervisor of UK cohort while A.H. and A.K.A.K. managed the Bangladeshi cohort studies. B.W.B. oversaw data collection and phenotyping of subjects in Bangladeshi cohorts. B.A.K. carried out sample collection and phenotyping in the EFSOCH cohort. I.D.M. provided technical support in DNA isolation and quality control analysis in Indian cohorts. S.S.N. and A.D. performed high throughput genotyping of Indian cohorts while B.O., Z.H. T.M.F. and R.M.F. were responsible for preparing samples and genotyping in the Bangladeshi and EFSOCH cohorts. S.S.N., A.D., A.S. cleaned Indian cohorts’ genotype data and generated imputed genotypes whereas R.N.B. performed QC and imputation of the Bangladeshi and EFSOCH cohort genotype data. A.R.W. defined the South Asian samples of the UK Biobank dataset using ancestry principal components. S.S.N. and R.N.B. performed the central analysis and wrote the first draft of the manuscript. All authors have contributed to manuscript writing, provided critical inputs and approved the final version of the manuscript.

## References

1. Barker DJ. Fetal origins of coronary heart disease. BMJ 311, 171–174 (1995).

2. Barker DJ, Gluckman PD, Godfrey KM, Harding JE, Owens JA, Robinson JS. Fetal nutrition and cardiovascular disease in adult life. Lancet 341, 938–941 (1993).

3. Godfrey KM, Barker DJ. Fetal nutrition and adult disease. Am J Clin Nutr 71, 1344S–1352S (2000).

4. Wells JC, Sharp G, Steer PJ, Leon DA. Paternal and maternal influences on differences in birth weight between Europeans and Indians born in the UK. PLoS One 8, e61116 (2013).

5. Fall CH, et al. Size at birth, maternal weight, and type 2 diabetes in South India. Diabet Med 15, 220–227 (1998).

6. Mi J, Law C, Zhang KL, Osmond C, Stein C, Barker D. Effects of infant birthweight and maternal body mass index in pregnancy on components of the insulin resistance syndrome in China. Ann Intern Med 132, 253–260 (2000).

7. Stein CE, Fall CH, Kumaran K, Osmond C, Cox V, Barker DJ. Fetal growth and coronary heart disease in south India. Lancet 348, 1269–1273 (1996).

8. Yajnik CS. Early life origins of insulin resistance and type 2 diabetes in India and other Asian countries. J Nutr 134, 205–210 (2004).

9. Ramachandran A, Snehalatha C, Viswanathan V, Viswanathan M, Haffner SM. Risk of noninsulin dependent diabetes mellitus conferred by obesity and central adiposity in different ethnic groups: a comparative analysis between Asian Indians, Mexican Americans and Whites. Diabetes Res Clin Pract 36, 121–125 (1997).

10. Beaumont RN, et al. Genome-wide association study of offspring birth weight in 86 577 women identifies five novel loci and highlights maternal genetic effects that are independent of fetal genetics. Hum Mol Genet 27, 742–756 (2018).

11. Freathy RM, et al. Variants in ADCY5 and near CCNL1 are associated with fetal growth and birth weight. Nat Genet 42, 430–435 (2010).

12. Horikoshi M, et al. Genome-wide associations for birth weight and correlations with adult disease. Nature 538, 248–252 (2016).

13. Horikoshi M, et al. New loci associated with birth weight identify genetic links between intrauterine growth and adult height and metabolism. Nat Genet 45, 76–82 (2013).

14. Warrington NM, et al. Maternal and fetal genetic effects on birth weight and their relevance to cardio-metabolic risk factors. Nat Genet 51, 804–814 (2019).

15. Hattersley AT, Tooke JE. The fetal insulin hypothesis: an alternative explanation of the association of low birthweight with diabetes and vascular disease. Lancet 353, 1789–1792 (1999).

16. Krishnaveni GV, Yajnik CS. Developmental origins of diabetes-an Indian perspective. Eur J Clin Nutr 71, 865–869 (2017).

17. Wells JC, Pomeroy E, Walimbe SR, Popkin BM, Yajnik CS. The Elevated Susceptibility to Diabetes in India: An Evolutionary Perspective. Front Public Health 4, 145 (2016).

18. Yajnik CS, et al. Neonatal anthropometry: the thin-fat Indian baby. The Pune Maternal Nutrition Study. Int J Obes Relat Metab Disord 27, 173–180 (2003).

19. Cousminer DL, Freathy RM. Genetics of early growth traits. Hum Mol Genet 29, R66–R72 (2020).

20. Hattersley AT, Beards F, Ballantyne E, Appleton M, Harvey R, Ellard S. Mutations in the glucokinase gene of the fetus result in reduced birth weight. Nat Genet 19, 268–270 (1998).

21. World Health Organization & United Nations Children’s Fund (UNICEF). Low Birthweight: Country, regional and global estimates. https://apps.who.int/iris/handle/10665/43184 (2004).

22. International Diabetes Federation. IDF report 7th edition. http://www.diabetesatlas.org (2015).

23. Prabhakaran D, Jeemon P, Roy A. Cardiovascular Diseases in India: Current Epidemiology and Future Directions. Circulation 133, 1605–1620 (2016).

24. Basu A, Sarkar-Roy N, Majumder PP. Genomic reconstruction of the history of extant populations of India reveals five distinct ancestral components and a complex structure. Proc Natl Acad Sci U S A 113, 1594–1599 (2016).

25. Reich D, Thangaraj K, Patterson N, Price AL, Singh L. Reconstructing Indian population history. Nature 461, 489–494 (2009).

26. Rao S, et al. Intake of micronutrient-rich foods in rural Indian mothers is associated with the size of their babies at birth: Pune Maternal Nutrition Study. J Nutr 131, 1217–1224 (2001).

27. Krishnaveni GV, Veena SR, Hill JC, Karat SC, Fall CH. Cohort profile: Mysore parthenon birth cohort. Int J Epidemiol 44, 28–36 (2015).

28. Carpenter MW, Coustan DR. Criteria for screening tests for gestational diabetes. Am J Obstet Gynecol 144, 768–773 (1982).

29. Potdar RD, et al. Improving women’s diet quality preconceptionally and during gestation: effects on birth weight and prevalence of low birth weight--a randomized controlled efficacy trial in India (Mumbai Maternal Nutrition Project). Am J Clin Nutr 100, 1257–1268 (2014).

30. Saffari A, et al. Effect of maternal preconceptional and pregnancy micronutrient interventions on children’s DNA methylation: Findings from the EMPHASIS study. Am J Clin Nutr 112, 1099–1113 (2020).

31. Krishna M, et al. Cohort Profile: The 1934-66 Mysore Birth Records Cohort in South India. Int J Epidemiol 44, 1833–1841 (2015).

32. Bhowmik B, et al. Maternal BMI and nutritional status in early pregnancy and its impact on neonatal outcomes at birth in Bangladesh. BMC Pregnancy Childbirth 19, 413 (2019).

33. Bhowmik B, et al. Vitamin D3 and B12 supplementation in pregnancy. Diabetes Res Clin Pract 174, 108728 (2021).

34. Knight B, Shields BM, Hattersley AT. The Exeter Family Study of Childhood Health (EFSOCH): study protocol and methodology. Paediatr Perinat Epidemiol 20, 172–179 (2006).

35. Hughes AE, et al. Fetal Genotype and Maternal Glucose Have Independent and Additive Effects on Birth Weight. Diabetes 67, 1024–1029 (2018).

36. Bycroft C, et al. The UK Biobank resource with deep phenotyping and genomic data. Nature 562, 203–209 (2018).

37. Genomes Project C, et al. A global reference for human genetic variation. Nature 526, 68–74 (2015).

38. McCarthy S, et al. A reference panel of 64,976 haplotypes for genotype imputation. Nat Genet 48, 1279–1283 (2016).

