## Supplemental figure 1 for "Associations of genetic scores for birth weight with newborn size and later Anthropometric traits and cardiometabolic risk markers in South Asians"

**A.**

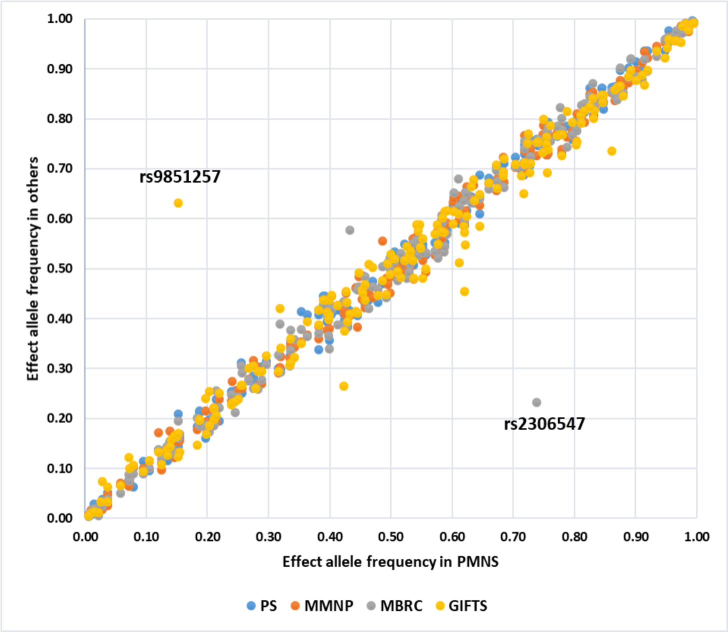


**B.**

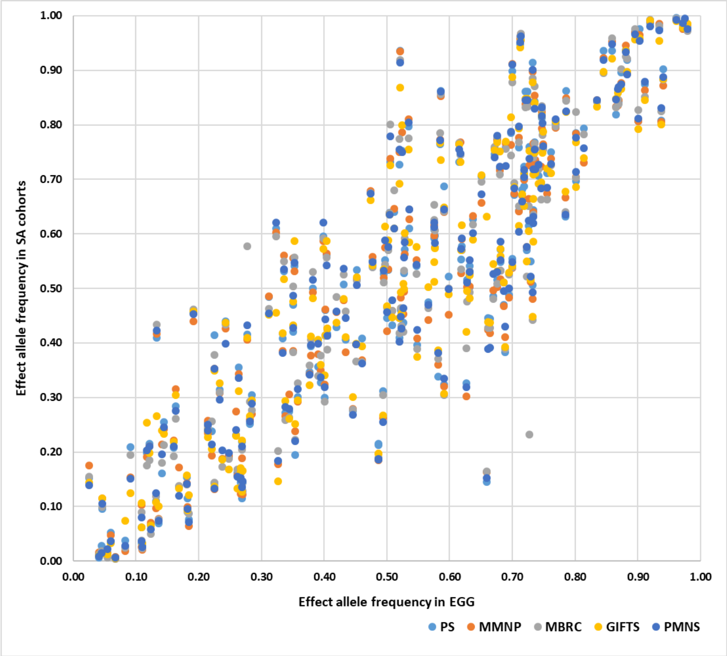


Supplementary figure 1: Comparison of the effect allele frequency of 196 birthweight-associated single nucleotide polymorphisms between EGG/UKBB and cohorts from South Asia (PMNS, Pune Maternal Nutrition Study; PS, Parthenon Study; MMNP, Mumbai Maternal Nutrition Project; MBRC, Mysore Birth Records Cohort; GIFTS, Bangladeshi Cohorts). (A) Between South Asian cohorts. PMNS is on the X-axis and the other South Asian cohorts are on the Y-axis, each marked with specific colours. The variants rs9851257 and rs2306547 are outliers in GIFTS and MBRC cohorts respectively. (B) Between EGG/UKBB and South Asians. EGG/UKBB is on the X-axis and the South Asian cohorts are on the Y-axis, each indicated by specific colours. EGG, Early Growth Genetics Consortium; UKBB, UK Biobank.
