## Supplemental tables 1-7 for "Associations of genetic scores for birth weight with newborn size and later Anthropometric traits and cardiometabolic risk markers in South Asians"

**Table S1A: Newborn and maternal anthropometry in the Indian cohorts**

| Traits | PMNS | | | PS | | | MMNP | | | MBRC | | |
|---|---|---|---|---|---|---|---|---|---|---|---|---|
| | Boys (N=271) | Girls (N=244) | All (N=515) | Boys (N=245) | Girls (N=266) | All (N=511) | Boys (N=271) | Girls (N=210) | All (N=481) | Boys (N=385) | Girls (N=299) | All (N=684) |
| Birthweight (kg) | 2.74 (0.33) | 2.62 (0.34) | 2.68 (0.34) | 2.96 (0.43) | 2.87 (0.38) | 2.91 (0.41) | 2.67 (0.37) | 2.59 (0.37) | 2.64 (0.37) | 2.81 (0.43) | 2.71 (0.39) | 2.76 (0.42) |
| Birth length (cm) | 48.2 (1.94) | 47.4 (1.92) | 47.8 (1.97) | 49.1 (2.13) | 48.6 (2.05) | 48.8 (2.11) | 48.5 (2.29) | 47.7 (2.14) | 48.2 (2.26) | 48.26 (3.00) | 47.7 (2.85) | 48.0 (2.95) |
| Ponderal index (kg/m$^3$) | 24.4 (2.17) | 24.6 (2.71) | 24.5 (2.44) | 24.9 (2.68) | 25.1 (2.82) | 25.0 (2.75) | 23.4 (2.35) | 23.9 (2.83) | 23.6 (2.58) | 25.3 (4.6) | 25.4 (5.16) | 25.3 (4.85) |
| Head circumference (cm) | 33.4 (1.18) | 32.7 (1.20) | 33.1 (1.24) | 34.2 (1.31) | 33.6 (1.19) | 33.9 (1.28) | 33.5 (1.20) | 32.9 (1.13) | 33.2 (1.21) | 33.7 (1.63) | 33.3 (1.50) | 33.6 (1.58) |
| Chest circumference (cm) | 31.4 (1.56) | 31.0 (1.59) | 31.2 (1.59) | 32.1 (1.68) | 32.0 (1.6) | 32.0 (1.64) | 31.0 (1.83) | 30.7 (1.69) | 30.9 (1.77) | NA | NA | NA |
| Abdominal circumference (cm) | 28.8 (1.93) | 28.7 (1.89) | 28.7 (1.91) | 30.0 (2.01) | 30.0 (1.83) | 30.0 (1.92) | 28.5 (2.07) | 28.4 (2.08) | 28.4 (2.07) | NA | NA | NA |
| Mid-upper arm circumference (cm) | 9.7 (0.87) | 9.6 (0.89) | 9.7 (0.88) | 10.4 (0.94) | 10.3 (0.89) | 10.4 (0.92) | 9.7 (0.80) | 9.7 (0.86) | 9.7 (0.82) | NA | NA | NA |
| Triceps skinfold (mm) | 4.2 (0.87) | 4.3 (0.87) | 4.3 (0.87) | 4.2 (0.91) | 4.3 (0.89) | 4.2 (0.90) | 4.1 (0.98) | 4.3 (1.11) | 4.2 (1.04) | NA | NA | NA |
| Subscapular skinfold (mm) | 4.2 (0.88) | 4.3 (0.91) | 4.2 (0.89) | 4.4 (0.89) | 4.6 (0.93) | 4.5 (0.91) | 4.0 (0.93) | 4.3 (1.03) | 4.2 (0.98) | NA | NA | NA |
| Gestational age (weeks) | 39.1 (1.05) | 39.0 (1.06) | 39.1 (1.06) | 39.4 (1.20) | 39.6 (1.07) | 39.5 (1.14) | 39.3 (1.18) | 39.3 (1.15) | 39.3 (1.17) | NA | NA | NA |
| Maternal age (years) | 21.4 (3.48) | 21.4 (3.65) | 21.4 (3.56) | 23.8 (4.16) | 23.8 (4.31) | 23.8 (4.24) | 24.7 (3.94) | 24.77 (3.77) | 24.73 (3.86) | NA | NA | NA |
| Maternal BMI (kg/m$^2$) | 18.2 (1.93) | 17.9 (1.87) | 18.1 (1.90) | 23.4 (3.51) | 23.8 (3.58) | 23.6 (3.55) | 20.2 (3.54) | 20.4 (3.86) | 20.3 (3.68) | NA | NA | NA |

The values are mean (SD). N, number of term babies with both genotype and phenotype data available; BMI, body mass index; SD, standard deviation; NA, not available. PMNS, Pune Maternal Nutrition Study; PS, Parthenon Study; MMNP, Mumbai Maternal Nutrition Project; MBRC, Mysore Birth Records Cohort.

**Table S1B: Newborn and maternal anthropometry in the Bangladeshi cohorts**

| Traits | Dhaka-WP2 | | | Dhaka-WP3 | | | UK-Bang | | |
|---|---|---|---|---|---|---|---|---|---|
| | **Boys (N=29)** | **Girls (N=24)** | **All (N=53)** | **Boys (N=162)** | **Girls (N=152)** | **All (N=314)** | **Boys (N=71)** | **Girls (N=72)** | **All (N=151)** |
| Birthweight (kg) | 2.99 (0.36) | 2.80 (0.39) | 2.90 (0.38) | 2.90 (0.39) | 2.77 (0.43) | 2.84 (0.42) | 3.18 (0.48) | 3.06 (0.41) | 3.12 (0.45) |
| Birth length (cm) | 46.5 (2.81) | 45.9 (2.23) | 46.2 (2.56) | 49.8 (2.58) | 49.4 (2.61) | 49.6 (2.60) | 47.1 (1.84) | 46.2 (2.08) | 46.7 (2.03) |
| Ponderal index ($kg/m^3$) | 29.9 (4.84) | 28.9 (3.89) | 29.5 (4.42) | 23.6 (3.50) | 23.0 (3.49) | 23.3 (3.50) | 28.7 (4.48) | 29.1 (4.21) | 28.9 (4.27) |
| Head circumference (cm) | 33.7 (1.49) | 33.0 (1.18) | 33.4 (1.39) | 33.1 (3.01) | 32.7 (1.49) | 32.9 (2.40) | 34.0 (1.08) | 33.1 (1.39) | 33.6 (1.31) |
| Chest circumference (cm) | NA | NA | NA | NA | NA | NA | 33.55 (2.37) | 33.25 (1.46) | 33.4 (1.97) |
| Abdominal circumference (cm) | NA | NA | NA | NA | NA | NA | 31.86 (2.60) | 30.95 (2.47) | 31.41 (2.56) |
| Mid-upper arm circumference (cm) | 10.0 (0.68) | 9.7 (0.74) | 9.9 (0.71) | 10.5 (2.76) | 10.0 (0.94) | 10.2 (2.09) | 11.4 (2.16) | 10.3 (1.96) | 10.9 (2.13) |
| Triceps skinfold (mm) | NA | NA | NA | NA | NA | NA | 4.8 (2.08) | 5.2 (1.80) | 5.0 (1.93) |
| Subscapular skinfold (mm) | NA | NA | NA | NA | NA | NA | 5.2 (2.02) | 5.3 (1.75) | 5.3 (1.87) |
| Gestational age (week) | 40.2 (0.86) | 40.4 (1.47) | 40.3 (1.17) | 39.2 (1.42) | 39.2 (1.63) | 39.2 (1.53) | 38.7 (4.61) | 39.2 (1.23) | 39.0 (3.44) |
| Maternal Age (years) | 20.07 (2.36) | 19.83 (2.66) | 19.91 (2.45) | 22.74 (4.00) | 22.66 (4.60) | 22.68 (4.29) | 29.22 (5.47) | 30.19 (5.31) | 29.68 (5.40) |
| Maternal BMI ($kg/m^2$) | 20.10 (3.36) | 21.36 (3.40) | 20.58 (3.40) | 22.39 (4.12) | 22.91 (3.92) | 22.65 (4.03) | 25.75 (4.02) | 26.79 (4.65) | 26.24 (4.34) |

The values are mean (SD). N, number of term babies with both genotype and phenotype data available; BMI, body mass index; SD, standard deviation; NA, not available; Dhaka-WP2, Work Package 2 of GIFTS; Dhaka-WP3, Work Package 3 of GIFTS; UK-Bang, London UK Bangladeshi cohort.

**Table S2A: Body size and composition, and cardiometabolic measures during childhood and early adolescence in the Indian cohorts**

| Traits | Childhood | | | Early adolescence | |
|---|---|---|---|---|---|
| | PMNS (N=608) | PS (N=562) | MMNP (N=696) | PMNS (N=604) | PS (N=516) |
| Age (years) | 6.17 (0.21) | 5.0 (0.04) | 5.85 (0.32) | 11.6 (0.93) | 13.53 (0.14) |
| Weight (kg) | 16.2 (1.9) | 15.2 (1.9) | 16.2 (2.5) | 29.3 (6.8) | 41.9 (8.6) |
| Height (cm) | 109.9 (4.7) | 105.6 (4.2) | 109.6 (4.9) | 139.6 (8.4) | 153.7 (6.9) |
| Body mass index (kg/m$^2$) | 13.4 (0.9) | 13.6 (1.1) | 13.4 (1.4) | 14.9 (2.1) | 17.7 (3.1) |
| Head circumference (cm) | 48.6 (1.5) | 48.5 (1.4) | 48.7 (1.5) | 51.3 (1.8) | 51.4 (1.4) |
| Waist circumference (cm) | 50.3 (2.6) | 45.9 (3.0) | 49.1 (3.6) | 57.4 (5.8) | 66.3 (7.9) |
| Mid-upper arm circumference (cm) | 15.2 (1.1) | 15.3 (1.2) | 15.4 (1.4) | 18.8 (3.1) | 22.1 (2.8) |
| Triceps skinfold (mm) | 6.3 (1.4) | 8.0 (2.1) | 7.4 (2.0) | 7.8 (3.3) | 13.3 (5.7) |
| Subscapular skinfold (mm) | 5.1 (1.1) | 6.2 (1.9) | 5.9 (1.7) | 6.9 (3.9) | 13.9 (7.1) |
| Fat percent (%) | 19.6 (5.5) | 25.5 (5.5) | 15.3 (5.2) | 16.7 (6.6) | 21.7 (7.5) |
| Systolic blood pressure (mm) | 90.4 (12.2) | 96.6 (8.3) | 92.1 (8.5) | 106.3 (10.0) | 109.4 (8.1) |
| Diastolic blood pressure (mm) | 53.6 (10.2) | 58.1 (6.8) | 56.1 (7.6) | 62.6 (6.8) | 61.2 (7.0) |
| Fasting glucose (mg/dL) | 88.9 (9.5) | 87.0 (9.4) | 86.4 (11.8) | 87.2 (7.2) | 90.2 (9.1) |
| 120 minutes glucose(mg/dL) | 99.6 (20.1) | 106.3 (17.7) | 84.4 (16.1) | NA | NA |
| Fasting insulin (mU/L) | 3.70 (2.48) | 4.16 (3.16) | 4.17 (5.18) | 5.89 (3.26) | 6.49 (4.18) |
| HOMA-IR | 0.82 (0.6) | 0.89 (0.7) | 0.63 (0.6) | 1.27 (0.8) | 1.69 (1.2) |
| Total cholesterol (mg/dL) | 127.8 (22.2) | 135.0 (26.6) | 145.4 (33.1) | 131.9 (22.4) | 136.4 (26.9) |
| LDL-cholesterol (mg/dL) | 73.0 (20.2) | 41.3 (12.5) | 89.8 (27.2) | 77.7 (19.1) | 80.5 (20.8) |
| HDL-cholesterol (mg/dL) | 42.2 (9.9) | 80.5 (39.2) | 39.8 (8.7) | 42.4 (9.0) | 41.5 (9.8) |
| Triglycerides (mg/dL) | 62.9 (22.9) | 57.0 (26.7) | 79.2 (30.6) | 58.9 (21.5) | 71.8 (37.1) |

Values are mean (SD). N, Number of individuals where both genotype and phenotype data are available (variable with different traits). PMNS, Pune Maternal Nutrition Study; PS, Parthenon Study; MMNP, Mumbai Maternal Nutrition Project; HOMA-IR, homeostasis model assessment of insulin resistance, LDL, low density lipoprotein; HDL, high density lipoprotein; NA, not available.

**Table S2B: Body size and composition, and cardiometabolic measures in the Indian adult cohorts**

| Traits | PMNS Mother (N=543) | PMNS Father (N=402) | PS Mother (N=525) | PS Father (N=499) | MMNP Mother (N=691) | MBRC (N=684) |
|---|---|---|---|---|---|---|
| Age (years) | 27.9 (3.5) | 39.4 (4.11) | 28.9 (4.3) | 36.4 (4.71) | 32.9 (4.48) | 62.2 (5.42) |
| Weight (kg) | 44.4 (6.8) | 59.9 (11.0) | 56.4 (11.1) | 67.1 (11.1) | 55.0 (11.4) | 66.56 (13.82) |
| Height (cm) | 153.0 (5.1) | 165.5 (6.1) | 154.5 (5.3) | 167.6 (6.3) | 152.3 (5.5) | 158.4 (9.7) |
| Body mass index (kg/m$^2$) | 18.9 (2.7) | 21.8 (3.6) | 23.6 (4.5) | 23.9 (3.6) | 23.7 (4.7) | 26.6 (5.3) |
| Head circumference (cm) | 53.0 (1.5) | 54.6 (1.6) | 52.4 (1.5) | 54.7 (1.6) | 52.4 (1.4) | 53.2 (1.7) |
| Waist circumference (cm) | 65.8 (7.2) | 80.1 (9.6) | 82.2 (11.8) | 86.2 (10.3) | 77.7 (11.4) | 93.0 (12.3) |
| Hip circumference (cm) | 85.5 (7.2) | 88.5 (6.9) | 92.4 (8.7) | 92.8 (7.3) | 87.8 (7.8) | 95.7 (11.2) |
| Waist to hip ratio | 0.77 (0.07) | 0.90 (0.07) | 0.89 (0.07) | 0.93 (0.06) | 69.7 (9.52) | 0.98 (0.11) |
| Mid upper arm circumference (cm) | 23.5 (2.4) | 26.4 (2.6) | 26.6 (3.6) | 28.6 (2.8) | 26.5 (3.9) | 29.5 (3.9) |
| Triceps skinfold (mm) | 9.7 (4.6) | 8.8 (4.3) | 23.0 (9.7) | 13.3 (5.6) | 18.5 (7.2) | 19.3 (7.7) |
| Subscapular skinfold (mm) | 12.8 (6.5) | 12.4 (6.0) | 31.0 (12.6) | 26.1 (11.3) | 27.8 (11.5) | 31.3 (9.8) |
| Systolic blood pressure (mmHg)* | 107.3 (9.6) | 110.6 (9.2) | 108.7 (11.2) | 116.9 (14.7) | 107.6 (12.2) | 127.0 (15.5) |
| Diastolic blood pressure (mmHg)* | 63.7 (6.9) | 63.7 (8.1) | 65.7 (9.1) | 73.6 (11.1) | 67.2 (9.6) | 75.2 (10.7) |
| Fasting glucose (mg/dl)* | 94.2 (17.3) | 93.9 (18.8) | 100.2 (22.4) | 108.2 (36.5) | NA | 129.2 (52.4) |
| 120 minutes glucose (mg/dL) | 99.0 (29.1) | 93.9 (39.4) | 115.4 (51.1) | NA | NA | NA |
| Fasting insulin (mU/L)* | 5.00 (3.71) | 6.77 (4.78) | 8.72 (5.66) | 8.75 (5.65) | NA | 13.03 (13.18) |
| HOMA-IR* | 1.19 (0.93) | 1.63 (1.5) | 2.18 (1.6) | 2.45 (2.2) | NA | 4.03 (3.8) |
| Total cholesterol (mg/dl)* | 139.5 (27.6) | 155.5 (31.9) | 159.7 (29.9) | 176.9 (38.4) | NA | 181.4 (42.1) |
| LDL- cholesterol (mg/dl)* | 81.5 (23.6) | 96.1 (25.2) | 94.8 (24.0) | 102.6 (29.2) | NA | 107.7 (33.8) |
| HDL- cholesterol (mg/dl)* | 45.5 (11.4) | 40.4 (11.1) | 44.1 (9.1) | 38.5 (7.0) | NA | 44.6 (10.8) |
| Triglycerides (mg/dl)* | 63.3 (30.7) | 95.4 (56.4) | 104.2 (62.0) | 179.3 (121.7) | NA | 145.7 (76.5) |

Values are mean (SD). N, Number of individuals with genotype phenotype data available and this can be variable with different traits. BMI was additionally included as a covariate for analysis of traits marked with an asterisk (*). SD, standard deviation; PMNS, Pune Maternal Nutrition Study; PS, Parthenon Study; MMNP, Mumbai Maternal Nutrition Project; MBRC, Mysore Birth Records Cohort; HOMA-IR, homeostatic model assessment insulin resistance; LDL, low density lipoprotein; HDL, high density lipoprotein; NA, not available.

**Table S3A: Associations of fetal genetic score with birthweight in South Asian populations (excluding GDM mothers)**

| Cohort | fGS adjusted for sex and GA[@] | | | | fGS adjusted for sex, GA, and mGS[#] | | | |
|---|---|---|---|---|---|---|---|---|
| | N | Effect | SE | P | N | Effect | SE | P |
| PMNS | 512 | 0.009 | 0.004 | 0.048 | 441 | 0.010 | 0.005 | 0.047 |
| PS | 480 | 0.023 | 0.005 | $3.0x10^{-7}$ | 428 | 0.023 | 0.005 | $1.3x10^{-6}$ |
| MMNP* | 434 | 0.012 | 0.005 | 0.012 | 428 | 0.013 | 0.005 | 0.009 |
| Dhaka-WP2 | 40 | 0.010 | 0.009 | 0.286 | 40 | 0.009 | 0.008 | 0.266 |
| Dhaka-WP3 | 233 | 0.004 | 0.003 | 0.200 | 233 | 0.004 | 0.003 | 0.218 |
| UK-Bang | 75 | 0.012 | 0.006 | 0.058 | 75 | 0.011 | 0.007 | 0.094 |
| Meta-analysis | 1774 | 0.010 | 0.002 | $5.1x10^{-8}$ | 1645 | 0.010 | 0.002 | $1.8x10^{-7}$ |

***@, $I^2$= 64.90%, Het-P=0.014 and #, $I^2$=62.50%, Het-P=0.02***

**Table S3B: Associations of maternal genetic score with birthweight in South Asian populations (excluding GDM mothers)**

| Cohort | mGS adjusted for sex and GA[@] | | | | mGS adjusted for sex, GA, and fGS[#] | | | |
|---|---|---|---|---|---|---|---|---|
| | N | Effect | SE | P | N | Effect | SE | P |
| PMNS | 459 | 0.000 | 0.004 | 0.975 | 441 | 0.001 | 0.004 | 0.881 |
| PS | 444 | 0.010 | 0.004 | 0.031 | 428 | 0.010 | 0.004 | 0.020 |
| MMNP* | 428 | 0.000 | 0.004 | 0.946 | 428 | 0.001 | 0.004 | 0.874 |
| Dhaka-WP2 | 40 | 0.020 | 0.008 | 0.019 | 40 | 0.020 | 0.008 | 0.019 |
| Dhaka-WP3 | 233 | 0.004 | 0.003 | 0.256 | 233 | 0.003 | 0.003 | 0.280 |
| UK-Bang | 75 | 0.009 | 0.007 | 0.217 | 75 | 0.006 | 0.007 | 0.386 |
| Meta-analysis | 1679 | 0.005 | 0.002 | 0.014 | 1645 | 0.005 | 0.002 | 0.011 |

***@, $I^2$=34.30%, Het-P=0.179 and #, $I^2$=30%, Het-P=0.210***

Association analysis was performed using linear regression, with standardized birthweight adjusted for sex and gestational age as the dependent variable for each cohort separately, and finally the summary results were meta-analyzed. *In MMNP, the allocation group was additionally adjusted for. The effect size is in standard deviation units. The standard deviation of birthweight in kg in all the cohorts ranged between 0.34 to 0.45 kg. N, number of term babies; GA, gestational age; SE, standard error; $I^2$, heterogeneity; Het-P, P value for heterozygosity; P, P value; mGS, maternal genetic score; fGS, fetal genetic score; GDM; gestational diabetes mellitus; PMNS, Pune Maternal Nutrition Study; PS, Parthenon Study; MMNP, Mumbai Maternal Nutrition Project; Dhaka-WP2, Work Package 2 of GIFTS; Dhaka-WP3, Work Package 3 of GIFTS; UK-Bang, London UK Bangladeshi cohort.

**Table S4A: Country-wise meta-analysis of association of fetal genetic score with own birthweight**

| Cohort | fGS adjusted for sex and GA | | | | fGS adjusted for sex, GA and mGS | | | |
|---|---|---|---|---|---|---|---|---|
| | N | Effect | SE | P | N | Effect | SE | P |
| Indians | 2176 | 0.012 | 0.002 | $8.9x10^{-8}$ | 1361 | 0.015 | 0.003 | $6.4x10^{-8}$ |
| Bangladeshis | 517 | 0.017 | 0.004 | $1.4x10^{-4}$ | 517 | 0.015 | 0.004 | $5.5x10^{-4}$ |

**Table S4B: Country-wise meta-analysis of association of maternal genetic score with offspring birthweight**

| Cohort | mGS adjusted for sex and GA | | | | mGS adjusted for sex, GA and fGS | | | |
|---|---|---|---|---|---|---|---|---|
| | N | Effect | SE | P | N | Effect | SE | P |
| Indians | 1386 | 0.003 | 0.003 | 0.197 | 1361 | 0.004 | 0.0026 | 0.128 |
| Bangladeshis | 517 | 0.014 | 0.004 | $4.0x10^{-4}$ | 517 | 0.012 | 0.004 | 0.002 |

Association analysis was performed using linear regression with standardized birthweight adjusted for sex and gestational age as the dependent variable for each cohort independently and finally the summary results were meta-analyzed. In MMNP, the allocation group was additionally adjusted for. The effect size is in standard deviation units of birthweight per unit change in genetic score. The standard deviation of birthweight ranged from 0.34 to 0.45 kg. N, number of term babies having both genotype and phenotype date; SNP, single nucleotide polymorphism; GA, gestational age; SE, standard error; P, P value; fGS, fetal genetic score; mGS, maternal genetic score. Indians include Pune Maternal Nutrition Study (PMNS), Parthenon Study (PS), Mumbai Maternal Nutrition Project (MMNP) and Mysore Birth Records Cohort (MBRC). Bangladeshis include Work Package 2 of GIFTS (Dhaka-WP2), Work Package 3 of GIFTS (Dhaka-WP3), and UK-Bang, London UK Bangladeshi cohort.

**Table S5: Associations of fetal and maternal genetic scores with other birth measurements in South Asian populations**

| Trait | fGS adjusted for sex, GA and mGS | | | | | | | mGS adjusted for sex, GA and fGS | | | | | | |
|---|---|---|---|---|---|---|---|---|---|---|---|---|---|---|
| | N | Effect | L95 | U95 | P | $I^2$ | Het-P | N | Effect | L95 | U95 | P | $I^2$ | Het-P |
| Birth length | 1795 | 0.007 | 0.002 | 0.012 | 0.011 | 0 | 0.558 | 1795 | 0.003 | -0.002 | 0.008 | 0.222 | 37.5 | 0.156 |
| Ponderal Index | 1771 | 0.011 | 0.005 | 0.016 | $3.3x10^{-4}$ | 39.7 | 0.141 | 1771 | 0.001 | -0.004 | 0.006 | 0.713 | 0 | 0.441 |
| Head circumference | 1819 | 0.010 | 0.005 | 0.015 | $2.3x10^{-4}$ | 0 | 0.722 | 1819 | 0.002 | -0.002 | 0.007 | 0.320 | 0 | 0.970 |
| Chest circumference | 1452 | 0.013 | 0.007 | 0.018 | $2.8x10^{-6}$ | 12.0 | 0.333 | 1452 | 0.002 | -0.002 | 0.007 | 0.321 | 0 | 0.467 |
| Abdominal circumference | 1452 | 0.015 | 0.010 | 0.021 | $6.9x10^{-8}$ | 52.1 | 0.099 | 1452 | 0.002 | -0.003 | 0.007 | 0.463 | 52.2 | 0.099 |
| Mid-upper arm circumference | 1819 | 0.014 | 0.009 | 0.020 | $2.5x10^{-7}$ | 0 | 0.575 | 1819 | 0.005 | 0.000 | 0.010 | 0.034 | 0 | 0.992 |
| Triceps skinfold | 1430 | 0.013 | 0.007 | 0.018 | $1.6x10^{-5}$ | 11.8 | 0.334 | 1430 | 0.004 | -0.001 | 0.009 | 0.161 | 55.4 | 0.081 |
| Subscapular skinfold | 1429 | 0.012 | 0.007 | 0.018 | $2.4x10^{-5}$ | 0 | 0.518 | 1429 | 0.003 | -0.002 | 0.008 | 0.225 | 14.8 | 0.318 |

Association analysis was performed using linear regression with standardized birth measures adjusted for sex and gestational age as dependent variables, for each cohort independently and finally the summary results were meta-analyzed. In MMNP, the allocation group was additionally adjusted for, and in MBRC only sex was adjusted for, since gestational data was not available for the majority of the sample. The effect size is in standard deviation units. The South Asian populations include (from India) the PMNS, Pune Maternal Nutrition Study; PS, Parthenon Study; MMNP, Mumbai Maternal Nutrition Project; and MBRC, Mysore Birth Records Cohort; (from Bangladesh) WP2, Work Package 2 of GIFTS; Dhaka-WP3, Work Package 3 of GIFTS; and (from the UK) the UK-Bang, London UK Bangladeshi cohort. N, number of term babies having both genotype and phenotype date; SNP, single nucleotide polymorphism; GA, gestational age; L95 and U95, 95% confidence interval; $I^2$, heterogeneity; Het-P, P value for heterogeneity; P, P value; fGS, fetal genetic score; mGS, maternal genetic score.

**Table S6: Associations of birthweight with anthropometric and cardiometabolic traits in Indian children$ and adolescents#**

| Trait | Children | | | | | Adolescents | | | | |
|---|---|---|---|---|---|---|---|---|---|---|
| | N | Effect | P | $I^2$ | Het-P | N | Effect | P | $I^2$ | Het-P |
| Weight | 1674 | 0.849 | $1.9\times10^{-51}$ | 50.3 | 0.134 | 1081 | 0.590 | $9.2\times10^{-20}$ | 0 | 0.977 |
| Height | 1673 | 0.665 | $1.3\times10^{-31}$ | 46.8 | 0.153 | 1081 | 0.523 | $8.6\times10^{-17}$ | 0 | 0.901 |
| Body Mass Index | 1673 | 0.634 | $2.5\times10^{-27}$ | 0 | 0.681 | 1081 | 0.456 | $8.7\times10^{-11}$ | 0 | 0.828 |
| Head circumference | 1674 | 0.721 | $3.2\times10^{-39}$ | 44.7 | 0.164 | 1076 | 0.755 | $8.1\times10^{-27}$ | 0 | 0.348 |
| Waist circumference | 1672 | 0.661 | 8.6x10-$^{29}$ | 45.4 | 0.160 | 1059 | 0.462 | $1.7\times10^{-10}$ | 0 | 0.986 |
| Mid-upper arm circumference | 1674 | 0.558 | $7.1\times10^{-21}$ | 0 | 0.943 | 1075 | 0.406 | $5.9\times10^{-9}$ | 0 | 0.925 |
| Triceps skin-fold | 1673 | 0.245 | $3.0\times10^{-5}$ | 0 | 0.856 | 1075 | 0.321 | $5.7\times10^{-6}$ | 0 | 0.830 |
| Sub-scapular skin-fold | 1673 | 0.311 | $1.3\times10^{-7}$ | 0 | 0.800 | 1074 | 0.318 | $5.7\times10^{-6}$ | 0 | 0.657 |
| Fat percent | 1659 | 0.123 | 0.030 | 28.3 | 0.248 | 1048 | 0.145 | 0.031 | 0 | 0.516 |
| Systolic blood pressure* | 1657 | -0.140 | 0.024 | 4.7 | 0.350 | 1064 | -0.148 | 0.041 | 0 | 0.714 |
| Diastolic blood pressure* | 1658 | -0.162 | 0.010 | 52.2 | 0.123 | 1055 | -0.020 | 0.794 | 51.8 | 0.150 |
| Fasting glucose* | 1653 | -0.075 | 0.237 | 0 | 0.664 | 1071 | -0.042 | 0.580 | 0 | 0.959 |
| 120 minutes glucose* | 1624 | 0.045 | 0.478 | 35.7 | 0.211 | NA | NA | NA | NA | NA |
| Fasting insulin* | 1644 | 0.001 | 0.991 | 0 | 0.824 | 1072 | -0.055 | 0.405 | 0 | 0.627 |
| HOMA-IR* | 1570 | -0.025 | 0.691 | 0 | 0.838 | 1071 | -0.061 | 0.365 | 0 | 0.655 |
| Total cholesterol* | 1652 | -0.045 | 0.481 | 0 | 0.675 | 1072 | -0.059 | 0.441 | 68.9 | 0.073 |
| LDL-cholesterol* | 1652 | 0.030 | 0.638 | 0 | 0.742 | 1072 | -0.020 | 0.787 | 67.3 | 0.080 |
| HDL-cholesterol* | 1662 | -0.036 | 0.573 | 13.5 | 0.315 | 1072 | 0.075 | 0.318 | 0 | 0.414 |
| Triglycerides* | 1652 | -0.209 | $9.8\times10^{-4}$ | 41.3 | 0.182 | 1072 | -0.228 | 0.002 | 0 | 0.344 |

Association analysis was performed using linear regression with standardized log10 transformed traits as dependent variables for each cohort independently and finally the summary results were meta-analyzed. Age and sex were included as covariates in the regression model for all traits; BMI was additionally included as a covariate for analysis of traits marked with an asterisk (*). In MMNP, the allocation group was additionally adjusted for. $, The meta-analysis for children included those from the Pune Maternal Nutrition Study at 6 yrs, the Parthenon Study at 5 yrs and the Mumbai Maternal Nutrition Project at 7 yrs of age. #, Meta-analysis included adolescents from Pune Maternal Nutrition Study at 12 yrs and from Parthenon Study at 13.5 yrs; P, P value; $I^2$, heterogeneity; Het-P, P value for heterozygosity; HOMA-IR, homeostasis model assessment of insulin resistance; LDL, low density lipoprotein; HDL, high density lipoprotein; NA, not available. Those passing the Bonferroni corrected $P \leq 0.001$ were considered as statistically significant.

**Table S7: Meta-analysis of associations of maternal genetic score with anthropometric and cardiometabolic traits in Indian children$ & adolescents#**

| Traits | Children | | | | | Adolescents | | | | |
|---|---|---|---|---|---|---|---|---|---|---|
| | N | effect | P | $I^2$ | Het-P | N | effect | P | $I^2$ | Het-P |
| Weight | 1760 | 0.003 | 0.152 | 0 | 0.911 | 1028 | 0.002 | 0.523 | 0 | 0.611 |
| Height | 1759 | 0.002 | 0.409 | 0 | 0.505 | 1028 | 0.002 | 0.478 | 0 | 0.480 |
| Body mass index | 1759 | 0.003 | 0.255 | 0 | 0.654 | 1028 | 0.001 | 0.723 | 20.9 | 0.261 |
| Head circumference | 1760 | 0.001 | 0.808 | 0 | 0.977 | 1023 | 0.002 | 0.510 | 0 | 0.645 |
| Waist circumference | 1758 | 0.003 | 0.263 | 0 | 0.915 | 1005 | 0.001 | 0.758 | 0 | 0.398 |
| Mid-upper arm circumference | 1759 | 0.004 | 0.122 | 0 | 0.916 | 1023 | 0.003 | 0.353 | 0 | 0.407 |
| Triceps skinfold | 1759 | 0.003 | 0.263 | 0 | 0.550 | 1022 | -0.001 | 0.789 | 0 | 0.352 |
| Subscapular skinfold | 1759 | 0.003 | 0.227 | 0 | 0.628 | 1021 | 0.000 | 0.861 | 42.7 | 0.186 |
| Fat percent | 1753 | -0.001 | 0.661 | 3.5 | 0.355 | 993 | 0.000 | 0.909 | 0 | 0.449 |
| Systolic blood pressure* | 1740 | -0.002 | 0.348 | 0 | 0.967 | 1005 | -0.001 | 0.784 | 0 | 0.629 |
| Diastolic blood pressure* | 1741 | 0.000 | 0.891 | 0 | 0.954 | 1005 | -0.002 | 0.440 | 0 | 0.884 |
| Fasting glucose* | 1733 | -0.002 | 0.449 | 11.1 | 0.325 | 1018 | 0.000 | 0.994 | 0 | 0.826 |
| 120 minutes glucose* | 1710 | -0.002 | 0.413 | 0 | 0.767 | NA | NA | NA | NA | NA |
| Fasting insulin* | 1727 | 0.000 | 0.963 | 26.9 | 0.255 | 1019 | 0.001 | 0.696 | 0 | 0.746 |
| HOMA-IR* | 1662 | 0.000 | 0.838 | 52.6 | 0.121 | 1018 | 0.001 | 0.738 | 0 | 0.776 |
| Total cholesterol* | 1732 | 0.004 | 0.056 | 5.7 | 0.346 | 1019 | 0.002 | 0.422 | 0 | 0.752 |
| LDL-cholesterol* | 1733 | 0.003 | 0.209 | 46.8 | 0.153 | 1019 | 0.000 | 0.911 | 0 | 0.674 |
| HDL-cholesterol* | 1742 | 0.003 | 0.219 | 59.4 | 0.085 | 1019 | 0.005 | 0.094 | 0 | 0.385 |
| Triglycerides* | 1731 | 0.000 | 0.840 | 57 | 0.098 | 1019 | 0.001 | 0.761 | 18.0 | 0.269 |

Association analysis was performed using linear regression with standardized log10 transformed traits as the dependent variable for each cohort independently and finally the summary results were meta-analyzed. Age and sex were included as covariates in the regression model for all traits; BMI was additionally included as a covariate for analysis of traits marked with an asterisk (*). $, Meta-analysis for children included those from Pune Maternal Nutrition Study at 6 yrs, Parthenon Study at 5 yrs; Mumbai Maternal Nutrition Project at 7 yrs of age; #, Meta-analysis included adolescents from Pune Maternal Nutrition Study at 12 yrs and from Parthenon Study at 13.5 yrs; P, P value; $I^2$, heterogeneity; Het-P, P value for heterozygosity; SNP, single nucleotide polymorphism; HOMA-IR, homeostasis model assessment of insulin resistance; LDL, low density lipoprotein; HDL, high density lipoprotein; NA, not available. Those passing the Bonferroni corrected $P \leq 0.001$ were considered as statistically significant.
